# Travel-driven emergence and spread of SARS-CoV-2 lineage B.1.620 with multiple VOC-like mutations and deletions in Europe

**DOI:** 10.1101/2021.05.04.21256637

**Authors:** Gytis Dudas, Samuel L. Hong, Barney Potter, Sébastien Calvignac-Spencer, Frédéric S. Niatou-Singa, Thais B. Tombolomako, Terence Fuh-Neba, Ulrich Vickos, Markus Ulrich, Fabian H. Leendertz, Kamran Khan, Alexander Watts, Ingrida Olendraitė, Joost Snijder, Kim N. Wijnant, Alexandre M.J.J. Bonvin, Pascale Martres, Sylvie Behillil, Ahidjo Ayouba, Martin Foudi Maidadi, Dowbiss Meta Djomsi, Celestin Godwe, Christelle Butel, Aistis Šimaitis, Miglė Gabrielaitė, Monika Katėnaitė, Rimvydas Norvilas, Ligita Raugaitė, Rimvydas Jonikas, Inga Nasvytienė, Živilė Žemeckienė, Dovydas Gečys, Kamilė Tamušauskaitė, Milda Norkienė, Emilija Vasiliūnaitė, Danguolė Žiogienė, Albertas Timinskas, Marius Šukys, Mantas Šarauskas, Gediminas Alzbutas, Dovilė Juozapaitė, Daniel Naumovas, Arnoldas Pautienius, Astra Vitkauskienė, Rasa Ugenskienė, Alma Gedvilaitė, Darius Čereškevičius, Vaiva Lesauskaitė, Lukas Žemaitis, Laimonas Griškevičius, Guy Baele

## Abstract

Many high-income countries have met the SARS-CoV-2 pandemic with overwhelming sequencing resources and have identified numerous distinct lineages, including some with notably altered biology. Over a year into the pandemic following unprecedented reductions in worldwide human mobility, distinct introduced lineages of SARS-CoV-2 without sequenced antecedents are increasingly discovered in high-income countries as a result of ongoing SARS-CoV-2 genomic surveillance initiatives. We here describe one such SARS-CoV-2 lineage, carrying many mutations and deletions in the spike protein shared with widespread variants of concern (VOCs), including E484K, S477N and deletions HV69Δ, Y144Δ, and LLA241/243Δ. This lineage – designated B.1.620 – is known to circulate in Lithuania and has now been found in several European states, but also in increasing numbers in central Africa owing to important recent increases in genome sequencing efforts on the continent. We provide evidence of likely ongoing local transmission of B.1.620 in Lithuania, France, Germany, Spain, Belgium and the Central African Republic. We describe the suite of mutations this lineage carries, its potential to be resistant to neutralising antibodies, travel histories for a subset of the European cases, and evidence of local B.1.620 transmission in Europe. We make a case for the likely Central African origin of this lineage by providing travel records as well as the outcomes of carefully crafted phylogenetic and phylogeographic inference methodologies, the latter of which is able to exploit individual travel histories recorded for infected travellers having entered different European countries.

## INTRODUCTION

Over a year into the pandemic and with unprecedented reduction in human mobility worldwide, distinct SARS-CoV-2 lineages have arisen in multiple geographic areas around the world (Tegally et al., 2020; Hodcroft et al., 2021; Faria et al., 2021). New lineages are constantly appearing (and disappearing) all over the world and may be designated variant under investigation (VUI) if considered to have concerning epidemiological, immunological or pathogenic properties. So far, three lineages (i.e. B.1.1.7, B.1.351 and P.1 according to the Pango SARS-CoV-2 lineage nomenclature; Rambaut et al. (2020)) have been categorized as variants of concern (VOCs), owing to their profile in terms of their many mutations and/or deletions they carry, in combination with evidence of an increase in transmissibility, disease severity and possible reduced vaccine efficacy.

In some cases a lineage may rise to high frequency in one location and seed others in its vicinity, such as lineage B.1.177 that became prevalent in Spain and was later spread across the rest of Europe (Hodcroft et al., 2021). In others, reductions in human mobility, insufficient surveillance and passage of time allowed lineages to emerge and rise to high frequency in certain areas, as has happened with lineage A.23.1 in Uganda (Butera et al., 2021), a pattern reminiscent of holdover H1N1 lineages discovered in West Africa years after the 2009 pandemic (Nelson et al., 2014). In the absence of routine genomic surveillance at their origin location, diverged lineages may still be observed as travel cases or transmission chains sparked by such in countries that do have sequencing programmes in place. A unique SARS-CoV-2 variant found in Iran early in the pandemic was characterised in this way (Eden et al., 2020), and recently travellers returning from Tanzania (Oliveira et al., 2021) were found to be infected with a lineage bearing multiple amino acid changes of concern. As more countries launch their own SARS-CoV-2 sequencing programmes, introduced strains are easier to detect since they tend to be atypical of a host country’s endemic SARS-CoV-2 diversity, particularly so when introduced lineages have accumulated genetic diversity not observed previously, a phenomenon that is characterised by long branches in phylogenetic trees. In Rwanda, this was exemplified by detection of lineage B.1.380 (Butera et al., 2021) which was characteristic of Rwandan and Ugandan epidemics at the time. The same sequencing programme was then perfectly positioned to observe a sweep where B.1.380 was replaced by lineage A.23.1 (Butera et al., 2021), which was first detected in Uganda (Bugembe et al., 2021), and to detect the country’s first cases of B.1.1.7 and B.1.351. Similarly, sequencing programmes in Europe were witness to rapid displacement of pan-European and endemic lineages with VOCs, primarily B.1.1.7 (*e*.*g*. Lyngse et al. (2021)).

Given the appearance of VOCs towards the end of 2020 and continued detection of previously unobserved SARS-CoV-2 diversity, it stands to reason that more variants of interest (VOIs), and perhaps even VOCs, can and likely do circulate in areas of the world where access to genome sequencing is not available nor provided as a service by international organisations. Lineage A.23.1 (Bugembe et al., 2021) from Uganda and provisionally designated variant of interest A.VOI.V2 (Oliveira et al., 2021) from Tanzania might represent the first detections of a much more diverse pool of variants circulating in Africa. We here describe a similar case in the form of a lineage designated B.1.620 that first caught our attention as a result of what was initially a small outbreak caused by a distinct and diverged lineage previously not detected in Lithuania, bearing multiple VOC-like mutations and deletions, many of which substantially alter the spike protein.

The first samples of B.1.620 in Lithuania were redirected to sequencing because they were flagged by occasional E484K repeat PCR testing carried out on PCR-positive samples. Starting April 2nd 2021, targeted E484K PCR confirmed a growing cluster of cases with this mutation in Anykščiai municipality in Utena county with a total of 43 E484K+ cases out of 81 tested by April 28th (Figure S1). Up to this point, the Lithuanian genomic surveillance programme had sequenced over 10% of PCR-positive SARS-CoV-2 cases in Lithuania and identified few lineages with E484K circulating in Lithuania. To date, the only other E484K-bearing lineages in Lithuania had been B.1.351 (one isolated case in Kaunas county, and 12 cases from a transmission chain centered in Vilnius county) and B.1.1.318 (one isolated case in Alytus county), none of which had been found in Utena county despite a high epidemic sequencing coverage in Lithuania (Figure S2).

An in-depth search for relatives of this lineage on GISAID (Shu and McCauley, 2017) uncovered a few genomes from Europe initially, though more continue to be found since B.1.620 received its Pango lineage designation which was subsequently integrated into GISAID. This lineage now includes genomes from a number of European countries such as France, Switzerland, Belgium, Germany, England, Spain, The Netherlands, Ireland, and Portugal, as well as individual genomes from the United States (US). Interestingly, a considerable proportion of European cases turned out to be travellers returning from Cameroon. Since late April 2021, sequencing teams operating in central Africa, primarily working on samples from Central African Republic, Equatorial Guinea and Democratic Republic of the Congo have been submitting B.1.620 genomes to GISAID.

We here describe the mutations and deletions the B.1.620 lineage carries, many of which were previously observed in individual VOCs, but not in combination, and present evidence that this lineage likely originated in Cameroon and is likely to circulate in neighbouring countries in Central Africa where its prevalence is expected to be high. By combining collected travel records from infected patients entering different European countries, and by exploiting this information in a recently developed Bayesian phylogeographic inference methodology (Lemey et al., 2020), we reconstruct the dispersal of lineage B.1.620 from its inferred origin in Cameroon to several of its neighbouring countries, Europe and the US. Finally, we provide a description of local transmission in Lithuania, France, Spain and Germany through phylogenetic and phylogeographic analysis, and in Belgium through the collection of travel records.

## RESULTS

### B.1.620 carries numerous VOC mutations and deletions

Lineage B.1.620 attracted our attention due to large numbers of unique mutations (*i*.*e*. its genomes are distantly related to available references) of the Lithuanian genomes in nextclade analyses, and those genomes initially being assigned to clade 20A, corresponding to B.1 in Pangolin nomenclature (Rambaut et al., 2020). Meanwhile, Pangolin (using the 2021-04-01 version of pangoLEARN) variously misclassified B.1.620 genomes as B.1.177 or B.1.177.57 and occasionally as correct but unhelpful B.1, prior to official designation of B.1.620 by the Pango SARS-CoV-2 lineage nomenclature team. Closer inspection of B.1.620 genomes revealed that this lineage carries a number of mutations and deletions that have been previously observed individually in VOCs and VOIs (Figures 1 and S3), but had not been seen in combination. Despite sharing multiple mutations and deletions with known VOCs, lineage B.1.620 does not appear to be of recombinant origin (Figure S4).

**Figure 1.**
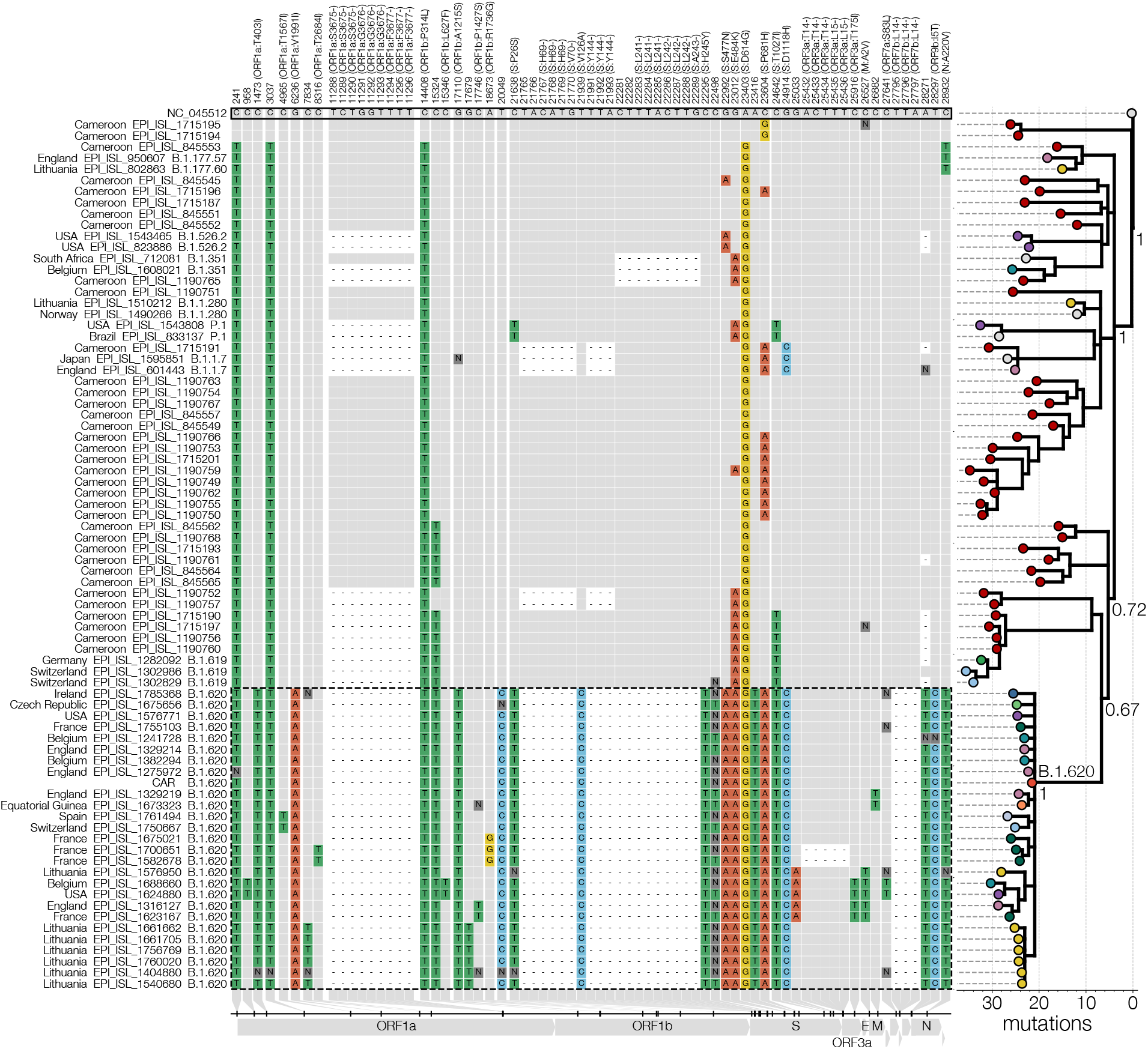
Lineage-defining SNPs of lineage B.1.620. Only SNPs that differentiate B.1.620 (genomes outlined with a dashed line) from the reference and are shared by at least two B.1.620 genomes are shown in the condensed SNP alignment. Sites identical to the reference (GenBank accession NC 045512) are shown in grey, changes from the reference are indicated and coloured by nucleotide (green for thymidine, red for adenosine, blue for cytosine, yellow for guanine, dark grey for ambiguities, black for gaps). If a mutation results in an amino acid change, the column label indicates the gene, reference amino acid, amino acid site, and amino acid change in brackets. The Bayesian phylogeny (branch lengths number of mutations) on the right shows the relationships between depicted genomes and was rooted on the reference sequence. Posterior probabilities of nodes leading up to lineage B.1.620 are shown near each node.

Through travel-related cases of B.1.620 discussed later we suspected that Cameroon is the immediate source of this lineage and therefore sought to identify close relatives of this lineage there. While genomic surveillance in Cameroon has been limited, the genomes that have been shared on GISAID are quite diverse and informative. A handful appear to bear several mutations in common with lineage B.1.620 and could be its distant relatives (Figure 1). Synonymous mutations at site 15324 and S:T1027I appear to be some of the earliest mutations that occurred in the evolution of lineage B.1.620, both of which are found in at least one other lineage associated with Cameroon (B.1.619), followed by S:E484K which also appears in genomes closest to lineage B.1.620. Even though the closest genomes to B.1.620 were sequenced from samples collected in January and February, lineage B.1.620 has 23 changes (mutations and deletions) leading up to it compared to the reference.

### B.1.620 is likely to escape antibody-mediated immunity

Like most currently circulating variants, B.1.620 carries the D614G mutation, which enhances infectivity of SARS-CoV-2, likely through enhanced interactions with the ACE2 receptor by promoting the up-conformation of the receptor binding domain (RDB) (Yurkovetskiy et al., 2020). Furthermore, B.1.620 contains P26S, HV69/70Δ, V126A, Y144Δ, LLA241/243Δ and H245Y in the N-terminal domain (NTD) of the spike protein. The individual V126A and H245Y substitutions are still largely uncharacterized to the best of our knowledge, but might be counterparts to the R246I substitution in B.1.351, and the latter may interfere with a putative glycan binding pocket in the NTD (Buchanan et al., 2021). All other mutations of B.1.620 in the NTD result in partial loss of neutralization of convalescent serum and NTD directed monoclonal antibodies (Wang et al., 2021). This indicates that these mutations present in B.1.620 may have arisen as an escape to antibody-mediated immunity (Liu et al., 2021). The spike protein of B.1.620 also carries both S477N and E484K mutations in the RBD, but in contrast to other VOCs not the N501Y or K417 mutations. Like the mutations in the NTD, S477N and E484K individually enable broad escape from antibody-mediated immunity (Liu et al., 2021). Moreover, deep mutational scanning experiments have shown that these substitutions also increase the affinity of the RBD for the ACE2 receptor (Starr et al., 2020). Both S477N and E484K occur on the same flexible loop at the periphery of the RDB-ACE2 interface (Lan et al., 2020).

We have modelled the RBD-ACE2 interface with the S477N and E484K substitutions using refinement in HADDOCK 2.4 (van Zundert et al., 2016). These models show that both individual substitutions and their combination produce a favorable interaction with comparable scores and individual energy terms to the ancestral RBD (see Supplementary Figure S5). Whereas S477N may modulate the loop conformation (Singh et al., 2021), E484K may introduce new salt bridges with E35/E75 of ACE2. These results indicate that B.1.620 may escape antibody-mediated immunity while maintaining a favorable interaction with ACE2. The remaining mutations in the spike protein – P681H, T1027I and D1118H – are uncharacterised to the best of our knowledge. Of these, P681H is also located on the outer surface of the spike protein, directly preceding the multibasic S1/S2 furin cleavage site (Hoffmann et al., 2020). In contrast T1027I and D1118H are both buried in the trimerisation interface of the S2 subunit (Walls et al., 2020).

### Local transmission of B.1.620 in Europe

Local transmission of B.1.620 in Lithuania has been established as a result of monitoring the outbreak in Anykščiai municipality (Utena county, Lithuania) via sequencing and repeat PCR testing of SARS-CoV-2 positive samples for the presence of E484K and N501Y, as well as looking for S gene target failure (SGTF) caused by the HV69Δ deletion. Genotypes identical to those found initially in Vilnius and Utena counties were later identified by sequencing in Alytus, Kaunas and Marijampolė counties, indicating continued transmission of lineage B.1.620 in-country. Interestingly, a single case in Tauragė county, Lithuania, identified by sequencing was a traveller returning from France found to be infected with a different genotype than the main outbreak lineage in Lithuania without evidence of onward transmission via local contact tracing efforts or genomic surveillance.

In addition to an ongoing disseminated outbreak of B.1.620 in Lithuania, genomes of this lineage have been found elsewhere in Europe. Though derived from separate introductions from the one that sparked outbreaks in Lithuania, other European B.1.620 genomes appear to indicate ongoing transmission in Europe, with clearest evidence of this in Germany and France, where emerging clades are comprised of identical or nearly identical genotypes (Figure 2). Presenting evidence for local transmission in Europe, B.1.620 genomes from countries like Spain and Belgium (also see next section) were notably picked up by baseline surveillance and thus are likely to represent local circulation, though presumably at much lower levels at the time of writing. Figure 2 shows the aforementioned local transmission clusters in Lithuania, Spain (Vilassar De Mar, province of Barcelona), France (see below), and Germany (state of Bavaria).

**Figure 2.**
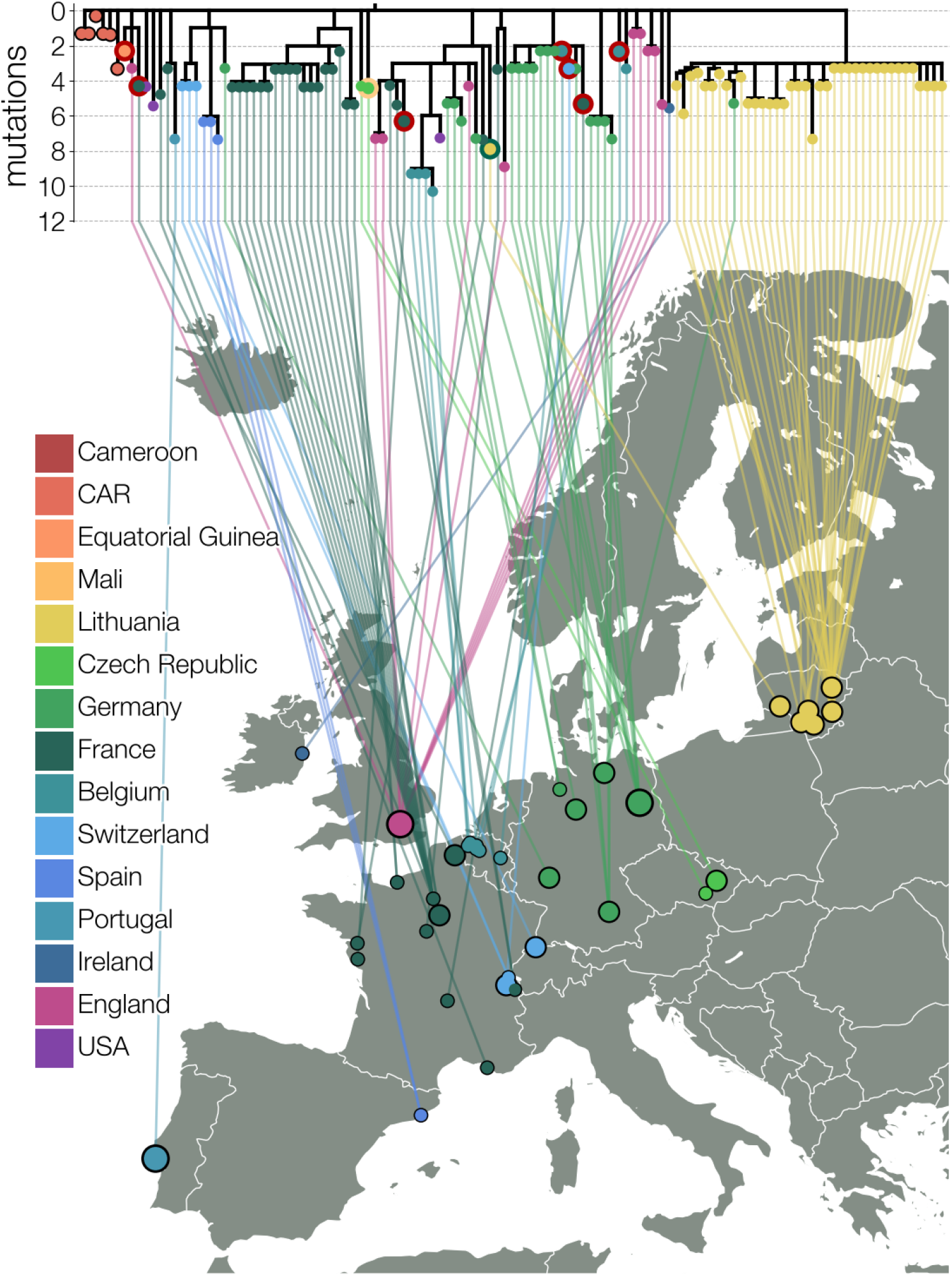
Maximum likelihood tree of lineage B.1.620 in Europe. Relationships between B.1.620 genomes, coloured by country of origin (same as Figure 1) with a thicker coloured outline indicating country of origin for travel cases. At least seven genomes shown (samples collected in Belgium, Switzerland, France, and Equatorial Guinea) are from individuals who returned from Cameroon, one is from a traveler returning from Mali and one Lithuanian case returned from France. Genomes from CAR and Czechia (returning traveler from Mali) are descended from the original B.1.620 genotype, while the genome from Equatorial Guinea is already closely related to genomes found in UK and happens to be a travel case from Cameroon. Each genome is connected to available geographic location in Europe with smallest circles indicating municipality-level precision, intermediate size corresponding to county level information (centered on county capital) and largest circle sizes indicating country level information (centered on country capital). Countries are assigned the same colours as in Figures 1 and 3.

In France, nine B.1.620 genomes (EPI ISL 1789089 – EPI ISL 1789097) were recently obtained from a large contact tracing investigation of a single transmission chain. These infections in the municipality of Pontoise (Val d’Oise department, to the northwest of Paris) occurred in adults (ages 24 to 38) who were all asymptomatic at the time of sampling. Additional infections in Pontoise outside of this cluster occurred in four adults (ages 29 to 57) and form a monophyletic cluster with the other nine infected individuals (Figure S4). The putative index case for these infections has yet to be determined through contact tracing at the time of writing but these cases clearly point to the B.1.620 lineage circulating in the Val d’Oise department. These infections seem to stem from local ongoing transmission in the Île-de-France region, clustering with two patients ages 1 (sample from a children’s hospital in Paris: Hôpital Necker-Enfants malades) and 69. These infections in Île-de-France in turn cluster with two infections from Le Havre (region of Normandy; 180km from Pontoise), pointing to either a travel event from Normandy to Île-de-France or possible local transmission in the north of France (Figure S4).

### B.1.620 likely circulates at high frequency in Central Africa

In the absence of routine surveillance at a location, sequencing infected travellers originating from there constitutes the next most efficient way to monitor distinct viral populations. This has been used successfully to uncover cryptic outbreaks of Zika virus in Cuba (Grubaugh et al., 2019) and SARS-CoV-2 in Iran at the beginning of the pandemic (Lemey et al., 2020). The latter study describes a novel approach to accommodate differences in sampling location and location of infection, and is hence specifically targeted to exploit recorded travel histories of infected individuals in Bayesian phylogeographic inference, rather than arbitrarily assigning the origin of the sample to either location. Of the collected B.1.620 genomes in our data set, seven are from travellers returning from Cameroon whereas six were sampled in the Central African Republic (CAR) near the border with Cameroon, indicating the most plausible geographic region where B.1.620 is circulating widely to be Central Africa (Figure S6). Given the limited sequencing capacity in these locations, we can only bound the ultimate origin of B.1.620 by neighbouring countries in Africa that have recently carried out sufficient sequencing to rule out high prevalence of B.1.620: Angola and South Africa to the south, Kenya to the east, and Togo with Nigeria to the northwest.

The collected individual travel histories themselves point to several independent introductions of B.1.620 into Europe, with documented cases of infected travelers returning from Cameroon to Belgium, France and Switzerland, and from Mali to Czechia (Figure 3). We note that the metadata for a returning traveler from Cameroon to Belgium (EPI ISL 1498300) presents evidence of ongoing local transmission within Belgium of B.1.620. Whereas this patient had spent time in Cameroon from the 16th of January until the 7th of February, a positive sample was only collected on the 15th of March, 2021. Even when assuming a lengthy infectious period of up to twenty days (Byrne et al., 2020), this patient’s infection can not stem from his prior travel to Cameroon, which indicates an infection with B.1.620 within Belgium and hence stemming from contact within the patient’s community. This is reinforced by a more recent sample from Belgium (EPI ISL 1620228), taken on the 28th of March, for which no travel history could be recorded and the patient declared not having left the country.

**Figure 3.**
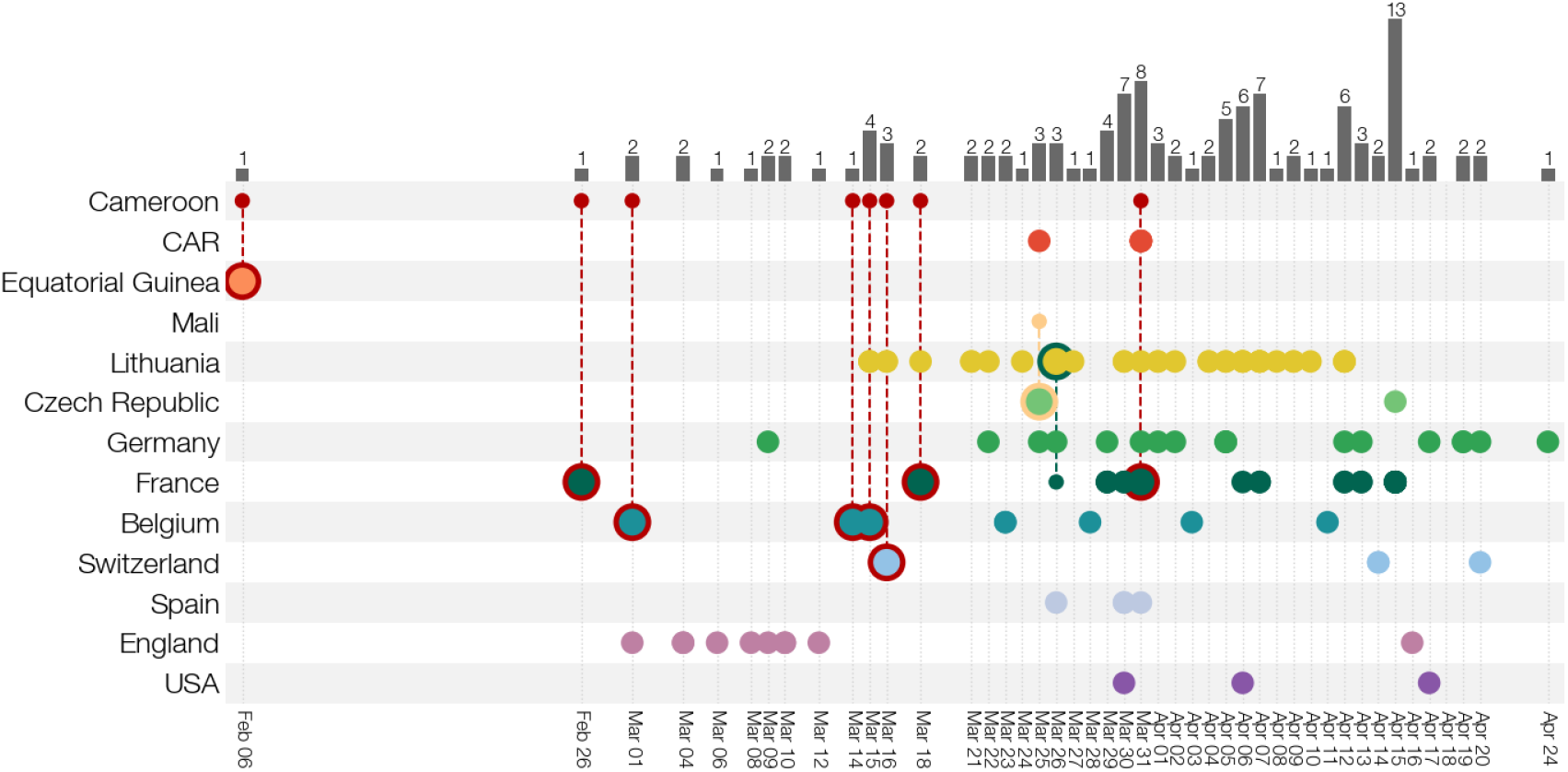
Known locations and travel history of B.1.620 cases. Collection dates of B.1.620 genomes are shown for each country (rows). Genomes from travellers are outlined with colour indicating travel of origin (e.g. red for Cameroon) and connected to a smaller dot indicating which country’s diversity is being sampled at travel destination. Bars at the top indicate the number of genomes of B.1.620 available for a given date across all countries. Countries are assigned the same colours as in Figure 1.

Using a Bayesian phylogeographic inference methodology that accommodates individual travel histories we were able to reconstruct location-annotated phylogenies at both the continent and country levels. Figure 4A shows the MCC tree of the continent-level phylogeographic analysis, which yields 75% posterior support for an African origin of lineage B.1.620. From this inferred African origin, the variant then spread to different European countries via multiple introductions, which is confirmed by our collection of travel history records for individuals returning to these countries. Subsequent country-level phylogeographic analysis on – shown in Figure 4B – points to Cameroon as the likely origin of this lineage, with posterior support of just over 99%. From Cameroon, the variant is estimated to have spread to two of its neighbouring countries, CAR and Equitorial Guinea, and to Europe via a series of introductions, confirming what was also observed in our recorded travel history records. Interestingly, two separate introductions into the USA are inferred to have occurred from Belgium, and a single Lithuanian case – a returning traveller from France – does not cluster with the cluster of remaining Lithuanian sequences, illustrative of at least two independent introductions of lineage B.1.620 into Lithuania. Figure 4B also shows four separate B.1.620 introduction events from Cameroon into England.

**Figure 4.**
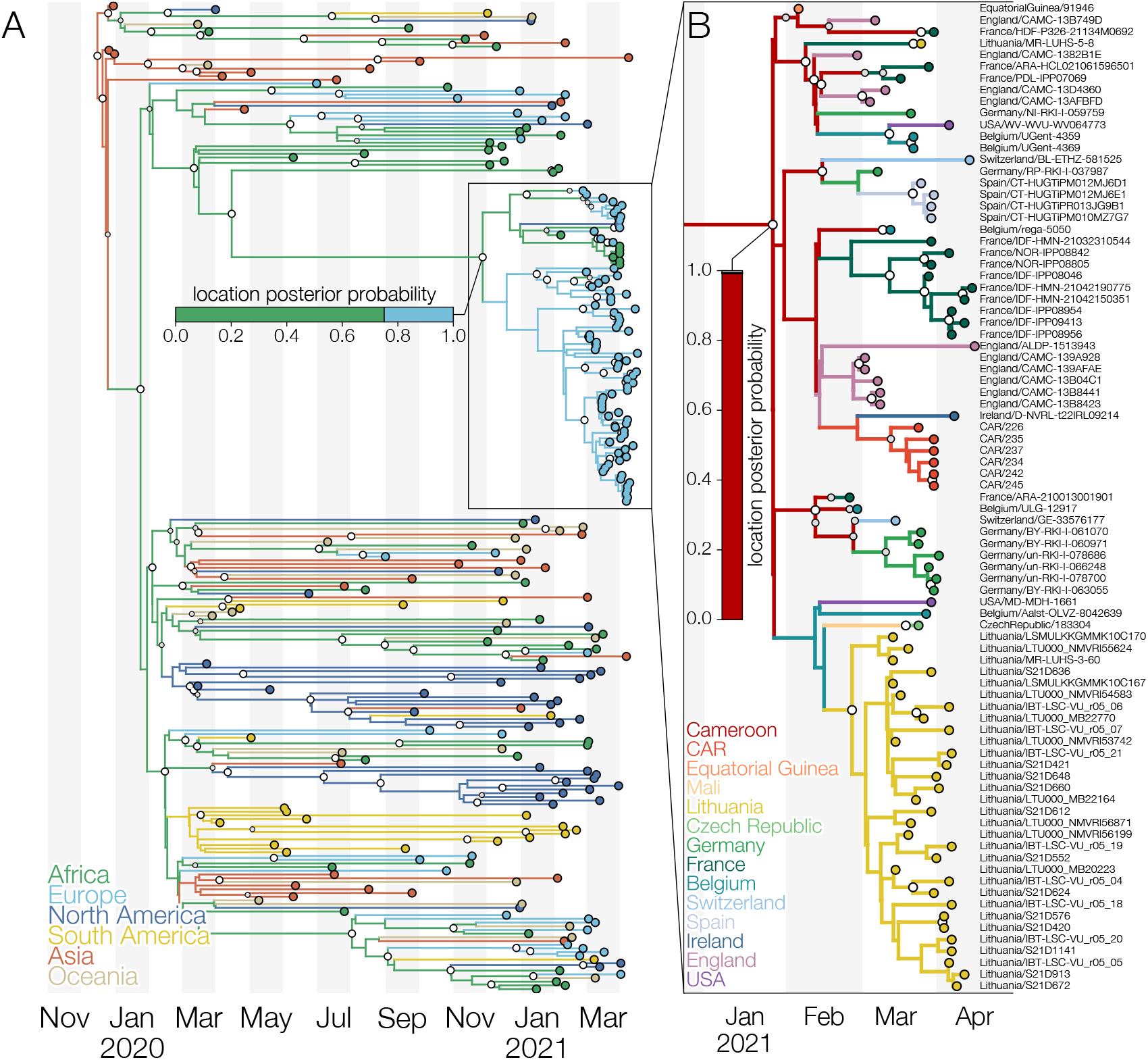
Maximum clade credibility trees of lineage B.1.620 coloured by reconstructed location. A) Global phylogeny of SARS-CoV-2 genomes with branches coloured by inferred continent from a Bayesian phylogeographic analysis that makes use of individual travel histories. Lineage B.1.620 is outlined and a horizontal bar shows posterior probability of its common ancestor existing in a given continent. Africa is reconstructed as the most likely location (posterior probability 0.75) where B.1.620 originated. B) Phylogeny of lineage B.1.620 and its closest relatives with branches coloured by inferred country from a Bayesian phylogeographic analysis that makes use of travel histories. A vertical bar shows posterior probabilities of where the common ancestor of B.1.620 existed. In this analysis Cameroon is reconstructed as the most likely location (posterior probability 0.99) of the common ancestor of lineage B.1.620. Larger white dots at nodes indicate nodes with posterior probability of at least 95%, while smaller grey circles indicate nodes with posterior probability of at least 50%.

Air passenger flux out of Cameroon (Figure 5) shows that many travellers had African countries as their destination, including many that have not reported any B.1.620 genomes to date. This suggests that B.1.620 should be widespread in Africa and its detection in Europe has mostly occurred in countries with recent active genomic surveillance programmes. Detections of B.1.620 in African states neighbouring Cameroon (Equatorial Guinea and DRC), even at low sequencing levels, suggests that B.1.620 may already be prevalent in central Africa. We find this apparent rise to high frequency and rapid spread across large areas of Africa noteworthy in light of other findings reported here, namely that currently available B.1.620 genomes appeared suddenly in February 2021 (Figure 3), are genetically homogeneous (Figure 2), and to date have no clear close relatives (Figure 1).

**Figure 5.**
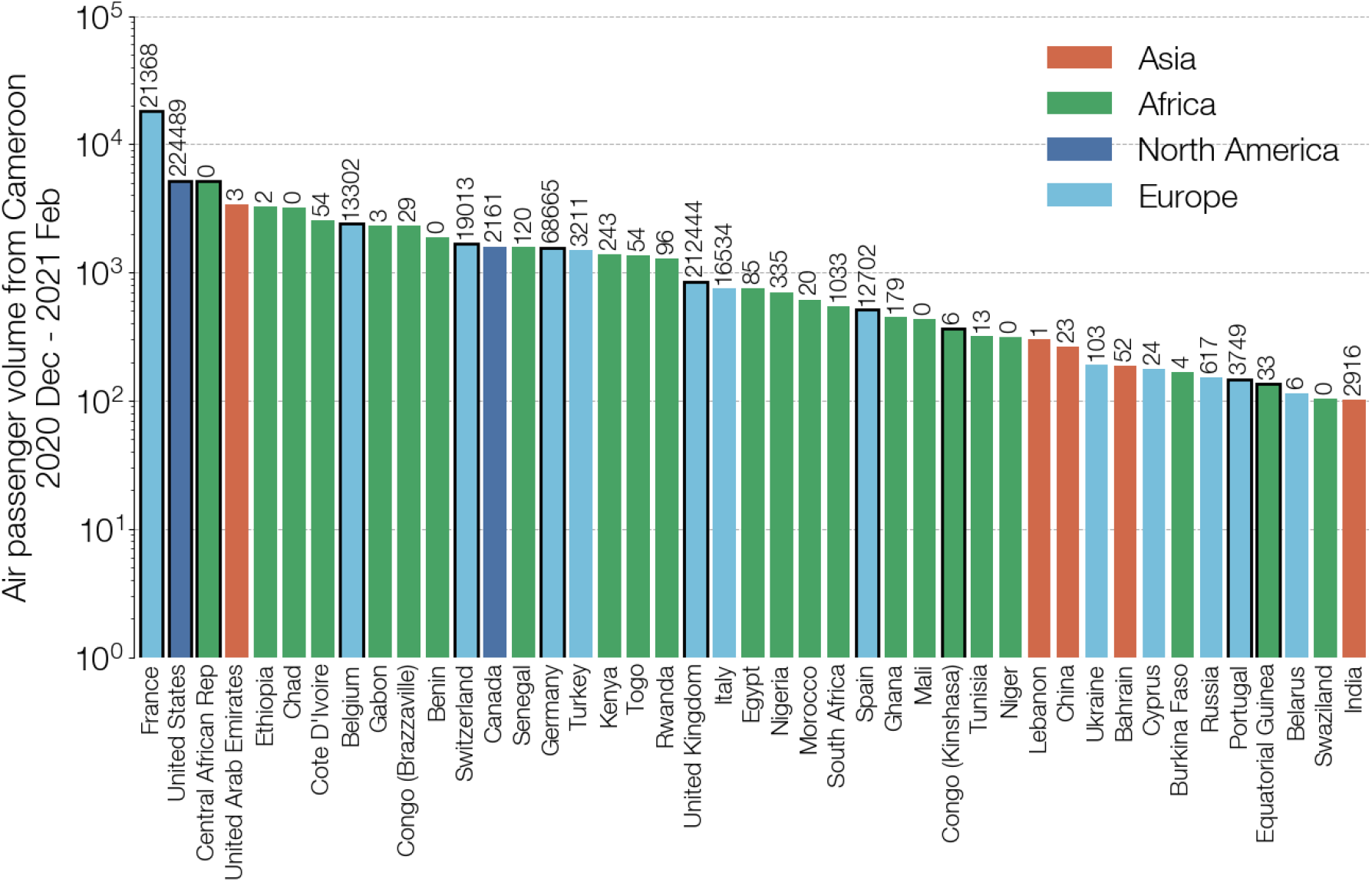
Total air passenger flows out of Cameroon between 2020 December and 2021 February. Destination countries are coloured by continent, sorted by total number of passengers, and limited to countries where at least 100 passengers have arrived from Cameroon between 2021 December and 2021 February. Numbers above each country’s bar indicate the total number of genomes on GISAID from that country since January 1st 2021, according to GISAID’s 2021-04-30 metadata release. Bars outlined in black represent countries that have submitted at least one B.1.620 genome as of 2021 May 04.

## DISCUSSION

In this study we have presented evidence that a SARS-CoV-2 lineage designated B.1.620, first detected in Europe in late February, is associated with Central Africa, where it appears to circulate at very high prevalence, and has been introduced into Europe and the US on multiple occasions. A fair number of known B.1.620 genomes that were sequenced in Europe stem from travel-related cases returning from Cameroon (Figure 3), suggesting that it is likely to be the immediate source of this lineage. SARS-CoV-2 genomes from CAR from near the border with Cameroon add weight to this hypothesis. So far the only observation that is difficult to explain is the Czech case returning from Mali, since Mali is over 1,000 km away from Cameroon. If B.1.620 were to be introduced to Mali via land routes it would first need to rise to high frequency in countries between Central Africa and Mali, at very least Nigeria and Niger. The lack of any B.1.620 genomes from Nigeria to date, despite higher air passenger volumes and active genomic surveillance in Nigeria (Figure 5) requires long distance travel to explain how B.1.620 reached Mali. Though possibly far-fetched, high prevalence of B.1.620 in Central Africa and the involvement of EU military both in CAR (EU, 2016) and in Mali (EU, 2013) may resolve this outlier.

In addition to the multiple introductions of the B.1.620 lineage we observe (Figure 3) and estimate (Figure 4) in Europe and North America, we also found evidence of local transmission of this lineage in Europe, with clearest evidence in Lithuania (Figure S1) followed by Germany and France (Figure 3), and finally Belgium and Catalonia, where B.1.620 genomes were picked up by baseline surveillance and infected individuals did not report having travelled abroad. B.1.620 is worrying for several reasons – its genomes are genetically homogeneous – as it appeared suddenly in February 2021 bearing a large number of VOC-like mutations and deletions in common with multiple VOCs (Figure S3), yet in the absence of any clear close relatives or sampled antecedents (Figure 1). The discovery of a novel lineage bearing many mutations of concern and with indications that they are introduced from locations where sequencing is not routine, is concerning and such occurrences may become an alarming norm.

The continued lack of genomic surveillance in multiple areas of the world, let alone equitable access to vaccines to drive transmission down, will continue to undermine efforts to control SARS-CoV-2 everywhere. Without the ability to identify unusual variants, to observe their evolution and learn from it, and to evaluate how vaccine-induced immunity protects against them, any response enacted by individual countries is reactive and, much like the process of evolution that generates variants of concern, short-sighted. The emergence of B.1.1.7 was unprecedented and has had a devastating impact on the state of the pandemic, so it is concerning that similar information gaps in global genomic surveillance still persist to this day. As an example we have shown that B.1.620 lacks intermediate relatives, resulting in a long branch that connects this lineage to the ancestral genotype of B.1. This could be the result of gradual but unsampled evolution, perhaps even far away from Cameroon and its neighbours, but it could have also happened due to unusual selection pressures in immunosuppressed individuals (Choi et al., 2020) which is hypothesised for lineage B.1.1.7. The long branch leading to B.1.620 also means that we cannot reconstruct the order of mutations that have occurred during the genesis of this lineage and therefore whether some amino acid changes have allowed others to happen by altering the fitness landscape via epistatic interactions (Kemp et al., 2021). Given the number of VOC-like mutations B.1.620 has, this is a significant loss.

Our work highlights that global inequalities, as far as infectious disease monitoring is concerned, have tangible impacts around the world and that until the SARS-CoV-2 pandemic is brought to heel everywhere, nowhere is safe for long. Additionally, we highlight the importance of collecting and sharing associated metadata with genome sequences, in particular regarding individual travel histories, as well as collection dates and locations, all of which are important to perform detailed phylogenetic and phylogeographic analysis. We only observed one single instance where a GISAID entry was accompanied by travel information and had to request such information for all the samples in our core data set by contacting each individual lab. Whereas many labs were quick to provide the requested information, we were certainly not able to retrieve all related individual travel histories. The scientific community therefore still faces the important task of reporting and sharing such critical metadata in a consistent manner, an aspect that has been brought to attention again during the ongoing pandemic (Oude Munnink et al., 2020; Gardner et al., 2021).

## MATERIALS AND METHODS

### Study design

This study was initiated upon detection of SARS-CoV-2 strains in Lithuania bearing Spike protein amino acid substitutions E484K, S477N and numerous B.1.1.7-like (HV69/70Δ and Y144Δ) and B.1.351-like (LLA241/243Δ) deletions, amongst others. In Lithuania repeat PCR testing of SARS-CoV-2 positive samples is occasionally carried out to detect N501Y, E484K and S gene target failure (SGTF) caused by the HV69Δ deletion. Upon detection of E484K-positive cases samples were redirected to sequencing. Initially identified cases of B.1.620 were mistakenly classified by pangolin as B.1.177 or B.1.177.57, while nextclade (Hadfield et al., 2018) assigned it to clade 20A rather than the expected 20E (EU1), while highlighting that B.1.620 sequences bore many unique mutations compared to the closest sequence. Searching GISAID for mutations E484K, S477N, and HV69/70Δ, which are found in numerous VOCs individually but not in combination, identified additional genomes that contained other mutations and deletions found in B.1.620.

We downloaded all available sequences of this lineage from GISAID on April 29, 2021, and identified members that clearly belonged to this lineage. Prior to official lineage designation B.1.620 most of its genomes could be identified by the presence of Spike protein E484K and S477N mutations and the HV69/70Δ deletion. Some of B.1.620 genomes were excluded from phylogenetic analyses because they were misassembled (*e*.*g*. hCoV-19/Belgium/UZA-UA-24912930/2021 is missing deletions characteristic of this lineage but has the mutations) or had too many ambiguous sites (*e*.*g*. hCoV-19/France/ARA-HCL021061598501/2021) but we recovered travel information about them regardless as this may prove useful to perform travel history-aware phylogeographic reconstruction (Lemey et al., 2020).

### SARS-CoV-2 whole genome sequencing

Every sample that tests positive for SARS-CoV-2 by PCR in Lithuania with Ct values *<* 30 may be redirected by the National Public Health Surveillance Laboratory to be sequenced by the European Centre for Disease Prevention and Control (ECDC), Vilnius University Hospital Santaros Klinikos (VUHSK), Hospital of Lithuanian University of Health Sciences Kaunas Clinics (HLUHSKC), Vilnius University Life Sciences Centre (VULSC) or Lithuanian University of Health Sciences (LUHS). Samples of this particular lineage were sequenced by ECDC using in-house protocols, infrastructure and assembly methods, VUHSK using Illumina COVIDSeq reagents, Illumina MiSeq platform, and assembled with covid-19-signal (Nasir et al., 2020), HLUHSKC using Twist SARS-CoV-2 Research Panel reagents, Illumina NextSeq550 platform, and assembled with V-pipe (Posada-Céspedes et al., 2021), LUHS using ARTIC protocol, Oxford Nanopore Technologies MinION platform, and assembled using ARTIC bioinformatics protocol for SARS-CoV-2, and VULSC using ARTIC V3 protocol combined with Invitrogen Collibri reagents, Illumina MiniSeq platform, Illumina DRAGEN COVID Lineage combined with an in-house BLAST v2.10.18-based assembly protocol. Samples from CAR were sequenced using the very same ARTIC V3 protocol as the Lithuanian University of Health Sciences (LUHS).

All SARS-CoV-2 genomes used here (except for CAR genomes) were downloaded from GISAID. A GISAID acknowledgment table containing all genome accession numbers is included with this study.

### Associated travel history

When available on GISAID as part of the uploaded metadata, we made use of this associated metadata information and contacted the submitting labs to determine precise travel dates. For all other cases, we retrieved individual travel histories by contacting the submitting labs – who then in turn contacted either the originating lab or the patient’s general practitioner – for any travel records they may have available. This resulted in travel itineraries for 10 patients, with 7 of these also containing detailed dates for the recorded travel. When a returning traveller visited multiple countries on the return trip, we included all visited countries as possible locations of infection by using an ambiguity code in the phylogeographic analysis (Lemey et al., 2020). The travel history information collected can be found in Supplementary Table S1. While we were able to retrieve travel history for a fair number of cases, this information is considered private information in certain countries and we were hence unable to retrieve such data for a subset of our sequences.

SARS-CoV-2 genomes from the United Kingdom (UK) make up a sizeable proportion of any phyloge-netic and phylogeographic analysis, given the impressive sequencing efforts by the COVID-19 Genomics UK Consortium. Given the lack of individual travel histories for the English B.1.620 genomes in our data set, we investigated the passenger volumes from all airports in Cameroon to all airports internationally, incorporating volumes from both direct and connecting flights between December 2020 and February 2021, from the International Air Transportation Association (IATA; International Air Transport Association (2021)). These passenger data cover the time frame of our estimated B.1.620 lineage since its origin (see Results), with the passenger volumes for February having become available at the time of writing as these data needs be retrieved and processed. These air passenger flux data reveal a very real possibility of missing travel histories from Cameroon for English B.1.620 cases, given that over 98% (*i*.*e*. 852 out of 867) of the passengers from Cameroon to the UK during this time frame had an English airport (London, Manchester or Birmingham) as their final destination. At the time of writing, Public Health England declined to comment on a Cameroonian origin for some of their B.1.620 infections.

### Modelling RBD-ACE2 interaction

We have modelled the RBD-ACE2 interface with the S477N and E484K substitutions using the final refinement step of HADDOCK 2.4 (van Zundert et al., 2016). We used the crystal structure of ACE2 (19-615) bound to SARS-CoV-2 RBD (PDB ID: 6m0j; Lan et al. (2020)) as a starting point and introduced the substitutions using UCSF ChimeraX (Goddard et al., 2018). We used default parameters for refinement with extended molecular dynamics (MD) simulation (steps for heating phase: 200, steps for 300K phase: 2500, steps for cooling phase: 1000).

### Phylogenetic and phylogeographic analysis

We combined 85 sequences belonging to lineage B.1.620 with sequences from lineages that have circulated in Lithuania at appreciable levels: B.1.1.7, B.1.1.280, B.1.177.60 and other VOCs that share mutations with lineage B.1.620: B.1.351, P.1 and B.1.526.2. We included high-quality Cameroonian sequences that were closest to lineage B.1.620 as well as the reference SARS-CoV-2 genome NC 045512. This resulted in a core set of 141 genomes, which is visualised in Figure S4, that serves as the starting point for our phylogenetic and phylogeographic analyses. This core set was subsequently combined with 150 randomly selected sequences from the nextstrain global analysis on April 29, 2021 (https://nextstrain.org/ncov/global; Hadfield et al. (2018)) to provide context for the B.1.620 analysis, plus an additional two reference sequences Wuhan/Hu-1/2019 and Wuhan/WH01/2019. We filtered these sequences based on metadata completeness and added an additional four Chinese sequences as well as eight non-Chinese sequences from Asia spanning both A and B lineages, in order to balance representation of different continents in our analyses. These sequences were aligned in MAFFT (FFT-NS-2 setting; Katoh and Standley (2013)) with insertions relative to reference removed, and 5’ and 3’ untranslated regions of the genome that were susceptible to sequencing and assembly error trimmed. We employed TempEst (Rambaut et al., 2016) to inspect the data set for any data quality issues that could result in an excess or shortage of private mutations in any sequences, or would point to assembly or any other type of sequencing issues.

We performed Bayesian model selection through (log) marginal likelihood estimation to determine the combination of substitution, molecular clock and coalescent models that best fits the data. To this end, we employed generalized stepping-stone sampling (GSS; Baele et al. (2016)) by running an initial Markov chain of 5 million iterations, followed by 50 path steps that each comprise 100,000 iterations, sampling every 500th iteration. We found that a combination of a non-parametric skygrid coalescent model (Gill et al., 2013), an uncorrelated relaxed clock model with underlying lognormal distribution (Drummond et al., 2006) and a general time-reversible substitution model with among-site rate variation (GTR+Γ_4_; Tavaré (1986); Yang (1994)) provided the optimal model fit to the data.

We subsequently performed a discrete Bayesian phylogeographic analysis in BEAST 1.10.5 (Suchard et al., 2018) using a recently developed model that is able to incorporate available individual travel history information associated with the collected samples (Lemey et al., 2020; Hong et al., 2021). Exploiting such information can yield more realistic reconstructions of virus spread, particularly when travelers from unsampled or under sampled locations are included to mitigate sampling bias. When the travel date for a sample could not be retrieved, we treated the time when the traveler started the journey as a random variable, and specified normal prior distributions over these random variables informed by an estimate of time of infection and truncated to be positive (back-in-time) relative to sampling date. As in previous work (Lemey et al., 2020), we used a mean of 10 days before sampling based on a mean incubation time of 5 days (Lauer et al., 2020b), a constant ascertainment period of 5 days between symptom onset and testing (Lauer et al., 2020a), and a standard deviation of 3 days to incorporate the uncertainty on the incubation time.

In our phylogeographic analysis, we made use of Bayesian stochastic search variable selection (BSSVS) to simultaneously determine which migration rates are zero depending on the evidence in the data and infer ancestral locations, in addition to providing a Bayes factor test to identify migration rates of significance (Lemey et al., 2009). We first performed a continent-level phylogeographic analyses by aggregating sampling locations as well as the individual travel histories that occurred between continents. To ensure consistent spatial reconstruction regardless of sampling, we fixed the root location of this tree to be in Asia – so as to match the known epidemiology of the COVID-19 pandemic – and assumed a monophyly constraint on the A and B lineages. Conditional on the results of this analysis, we performed a country-level analysis on the B.1.620 lineage and its parental lineage, in order to substantially reduce the computational burden and statistical complexity associated with having 87 sampling locations in a travel history-aware phylogeographic analysis. We made use of the following prior specifications for this analysis: a gamma (shape = 0.001; scale = 1000) prior on the skygrid precision parameter, Dirichlet(1.0, *K*) priors on all sets of frequencies (with *K* the number of categories), Gamma prior distributions (shape = rate = 1.0) on the unnormalized transition rates between locations (Lemey et al., 2009), a Poisson prior (country level: *λ* =28; continent level: *λ* =5) on the sum of non-zero transition rates between locations, a CTMC reference prior on the mean evolutionary rate and as well as on the overall (constant) diffusion rate (Ferreira and Suchard, 2008). In the country-level analysis, we assumed a normally distributed root height prior on the time of origin of B.1.620’s parental lineage, with mean on the 5th of February 2020 and standard deviation of 2 weeks, as derived from the corresponding internal node’s 95% highest posterior density interval in the preceding continent-level analysis. These phylogeographic analyses ran for a total of 125 (continent-level) and 200 million (country-level) iterations, with the Markov chain being sampled every 25,000th iteration, in order to reach an effective sample size (ESS) for all relevant parameters of at least 200, as determined by Tracer 1.7 (Rambaut et al., 2018). We used TreeAnnotator to construct a maximum clade credibility (MCC) tree and used baltic (https://github.com/evogytis/baltic) to visualise it.

Since more B.1.620 genomes are turning up every day, the latest dataset we were able to compile for Figures 2, 3, and S4 included 121 B.1.620 genomes. Most new genomes in this dataset compared to the core dataset described earlier were sequences from Germany and Lithuania. These 121 genomes were combined with previously described representatives of VOCs B.1.1.7, B.1.351, P.1 and B.1.526.2, other lineages circulating in Lithuania (B.1.177.60 and B.1.1.280), and high quality SARS-CoV-2 genomes from Cameroon to yield a dataset with 182 genomes total. A maximum likelihood phylogeny was inferred from this dataset using PhyML (Guindon et al., 2010) under the HKY+Γ_4_ model of nucleotide substitution (Hasegawa et al., 1985; Yang, 1994) which was then rooted on the reference sequence and shown in Figure S4. To occupy less space in Figure 1 the number of B.1.620 genomes was reduced down to a representative set of 27, and a phylogeny was inferred using MrBayes v3.2 (Ronquist et al., 2012) under the HKY+Γ_4_ model of nucleotide substitution (Hasegawa et al., 1985; Yang, 1994) and rooted on the reference sequence. MCMC was run for 2 million states, sampling every 1000th state and convergence confirmed by checking that effective sample sizes (ESSs) were above 200 for every parameter.

### Data availability

A list of GISAID accessions for genomes used here, as well as phylogenetic trees and scripts used to generate the figures are available at https://github.com/evogytis/B.1.620-in-Europe.

## Supporting information

GISAID acknowledgment table

## Data Availability

Data allowed to be distributed and scripts to reproduce analyses and figures are available at https://github.com/evogytis/B.1.620-in-Europe

## ACKNOWLEDGMENTS

We gratefully acknowledge the authors from originating laboratories responsible for obtaining the speci-mens, as well as submitting laboratories where the genome data were generated and shared via GISAID, on which this research is based. An acknowledgment table with accessions of SARS-CoV-2 genomes used here is included.

We thank all involved in the collection and processing of SARS-CoV-2 testing and genomic data, as well as associated metadata on individual travel histories. In particular we would like to thank Marc Noguera Julian, Elisa Martro Catala, Samuel Cordey, Piet Maes, Keith Durkin, Lize Cuypers, Lien Cattoir, Veerle Matheeussen, Vincent Enouf, Sylvie van der Werf, Etienne Simon-Lorie`re, Tobias Schindler, Vladimira Koudelakova, Gabriel Gonzalez, Ariane Düx, Yanthe Nobel, Livia Patrono, Justas Dapkūnas, and Andrew J. Tatem.

SLH acknowledges support from the Research Foundation - Flanders (“Fonds voor Wetenschappelijk Onderzoek - Vlaanderen,” G0D5117N). BP and GB acknowledges support from the Internal Fondsen KU Leuven/Internal Funds KU Leuven (Grant No. C14/18/094). GB acknowledges support from the Research Foundation - Flanders (“Fonds voor Wetenschappelijk Onderzoek - Vlaanderen,” G0E1420N, G098321N). FHL and TFN were supported by WWF and German Research Council’s grant LE1813/14-1 (Great Ape Health in Tropical Africa), Research in CAR took place under permit #098/MRSIT/DIRCAB/CB.20, granted to TT by the Ministery of Scientific Research and Technological Innovation. JS acknowledges funding from the Dutch Research Council NWO Gravitation 2013 BOO, Institute for Chemical Immunology (ICI; 024.002.009). AMJJB acknowledges the support of European Union Horizon 2020 projects BioExcel (823830) and EOSC-Hub (777536) projects. AA and CB acknowledge the support of the French National Research Institute for Sustainable Development (IRD).

---

**Table S1.** Individual travel histories collected for the core genomic data set analysed in this study. Importantly, documented travel cases from Cameroon to several European countries were retrieved from the labs that submitted the genomes to GISAID, often with detailed travel dates. Additionally, we were able to retrieve one travel case from Belgium to Lithuania (a truck driver passing through Germany and Poland on the way) through contact tracing, but without the accompanying genome sequence.

| Accession ID | Sampling location | Sampling date | Travel location | Travel date |
| --- | --- | --- | --- | --- |
| EPI_ISL_1241728 | Belgium | 2021-03-01 | Cameroon | 2021-02-26 |
| EPI_ISL_1369646 | Switzerland | 2021-03-16 | Cameroon | N/A |
| EPI_ISL_1382294 | Belgium | 2021-03-14 | Cameroon | 2021-03-11 |
| EPI_ISL_1406653 | France | 2021-02-26 | Cameroon | N/A |
| EPI_ISL_1495980 | France | 2021-03-18 | Cameroon | 2021-03-04 |
| EPI_ISL_1576950 | Lithuania | 2021-03-26 | France | 2021-03-23 |
| EPI_ISL_1671822 | France | 2021-03-31 | Cameroon | 2021-03-26 |
| EPI_ISL_1673323 | Equatorial Guinea | 2021-02-06 | Cameroon | 2021-02-01 |
| EPI_ISL_1675656 | Czech Republic | 2021-03-25 | Mali | 2021-03-20 |
| unsampled | Lithuania | 2021-02-26 | Belgium/Germany/Poland | N/A |

**Figure S1.**
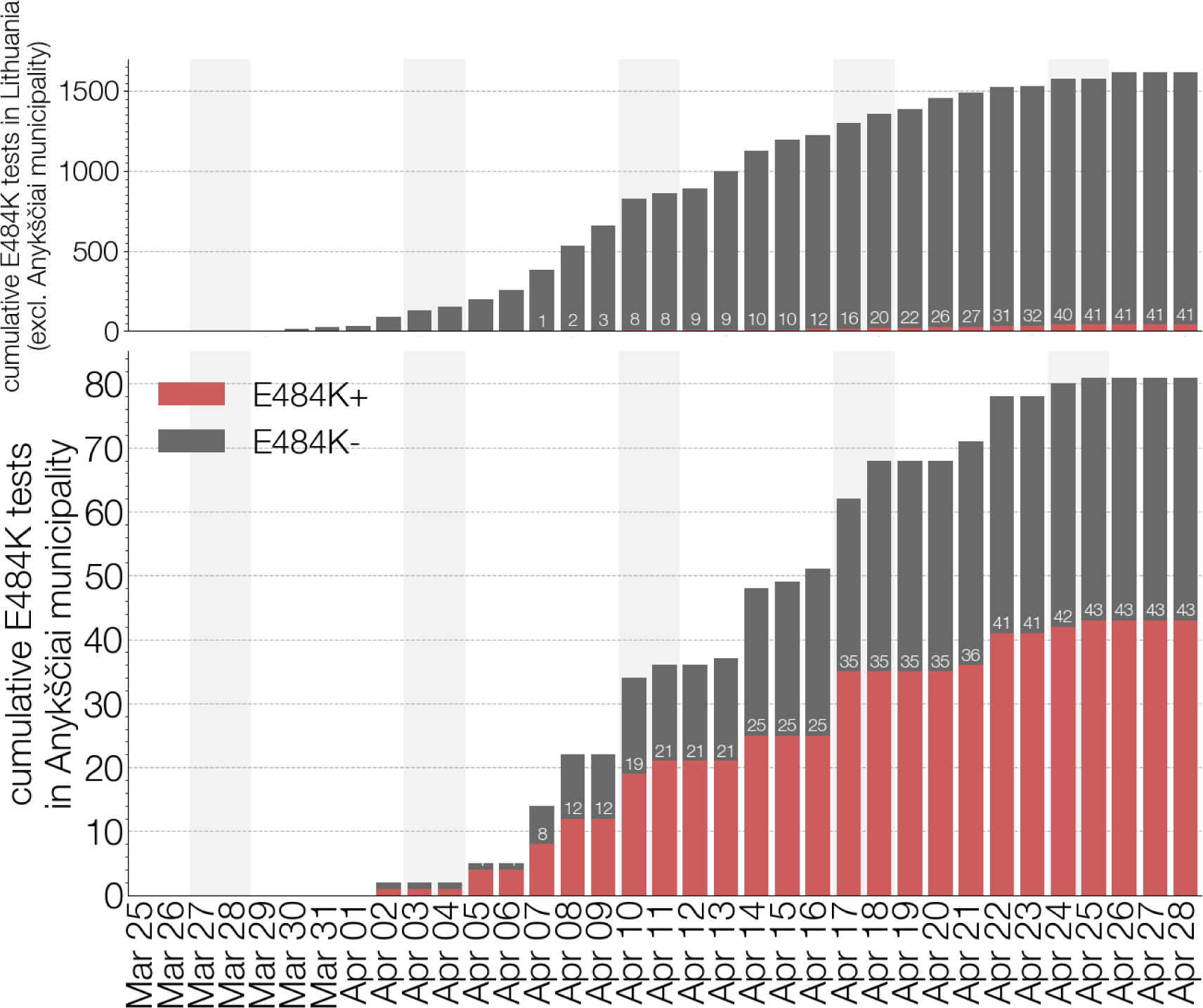
E484K PCR testing in Lithuania. Top panel shows the cumulative number of E484K PCR tests carried out in Lithuania (excluding Anyksčiai municipality) with E484K positive samples shown in red, and E484K negative samples shown in grey. Bottom panel shows the same information for the Anyksčiai municipality.

**Figure S2.**
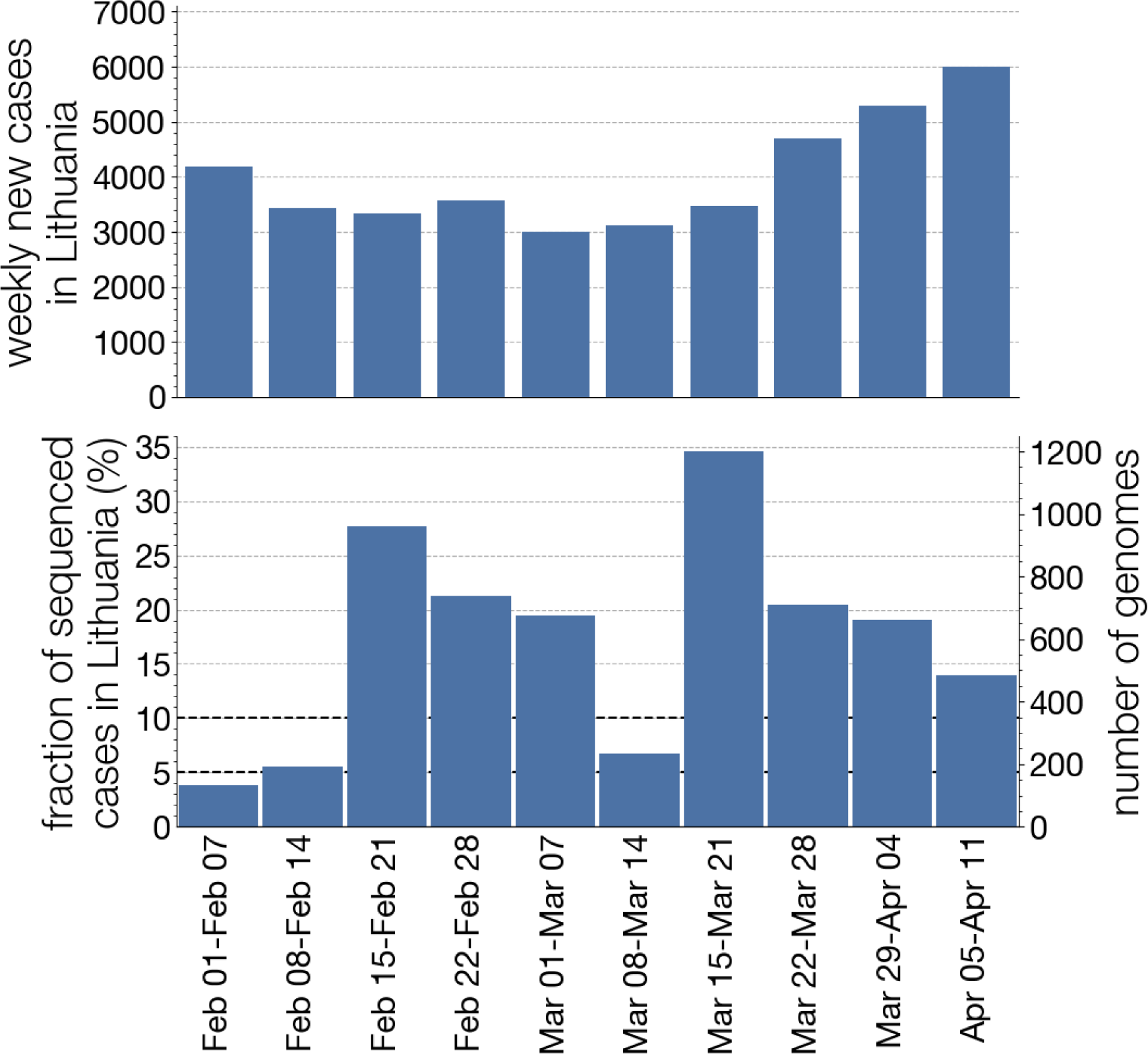
Lithuanian SARS-CoV-2 sequencing programme. Since its inception in February 2021, Lithuania’s SARS-CoV-2 sequencing programme has largely sequenced above the European Commission recommended 5% of positive cases. The sequencing programme began at the tail end of the second wave in Lithuania before restrictions were loosened in April 2021.

**Figure S3.**
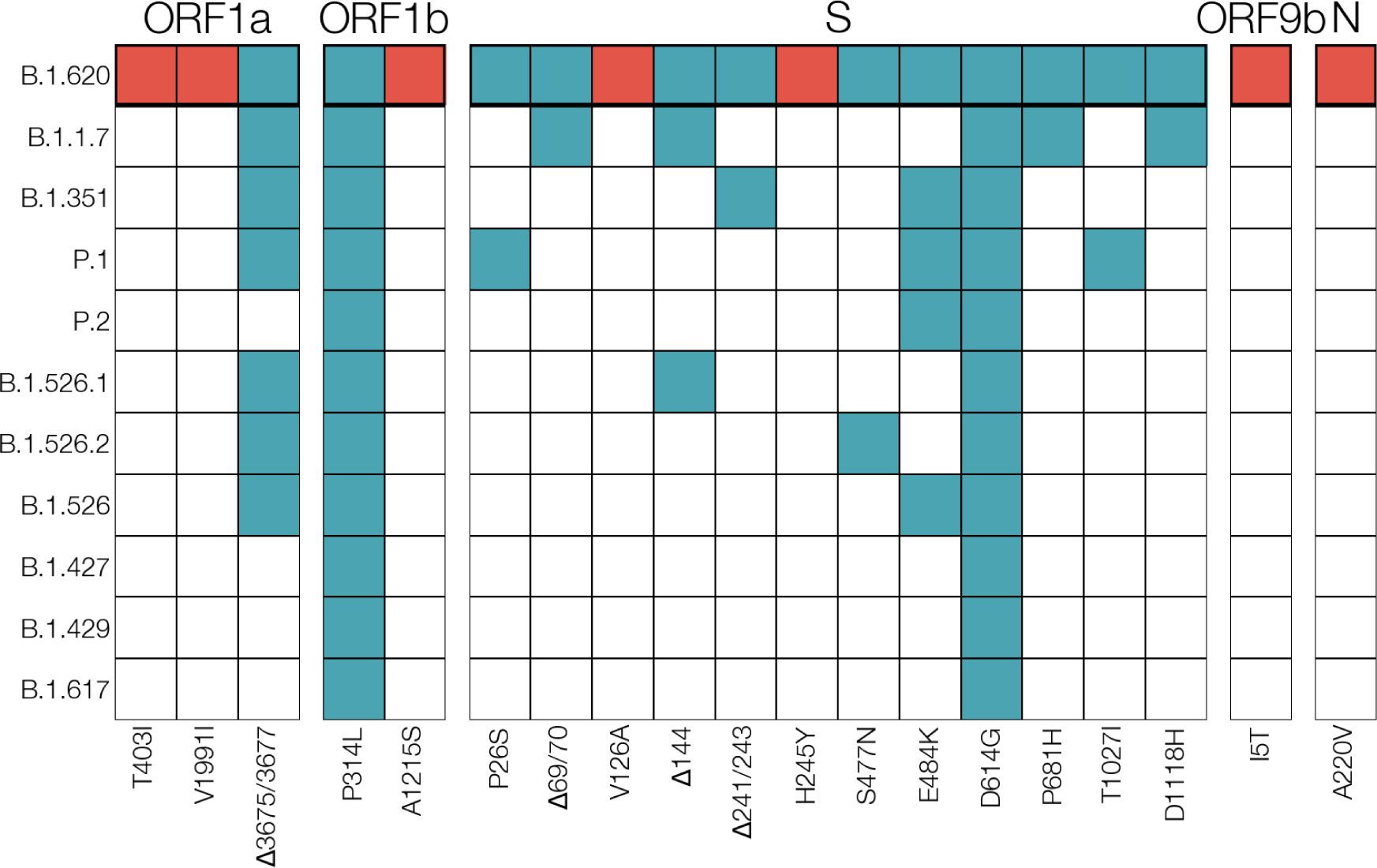
Comparison of amino acid mutations present in lineage B.1.620 and VOCs. Each row corresponds to lineage B.1.620 and current variants of concern (VOC) or interest (VOI). Each column is an amino acid change observed in lineage B.1.620 with cells indicating whether the mutation is unique to lineage B.1.620 in this comparison (red), shared with other VOCs/VOIs (blue) or absent (white). Of VOC amino acid changes lineage B.1.620 shares most in common with B.1.1.7 (ORF1a: SGF3675/3677Δ, S: Y144Δ, S: HV69/70Δ, S: P681H, and S: D1118H), followed by P.1 (ORF1a: SGF3675/3677Δ, S:P26S, S:E484K, S: T1027I) and B.1.351 (ORF1a: SGF3675/3677Δ, S: E484K, S: LLA241/243Δ).

**Figure S4.**
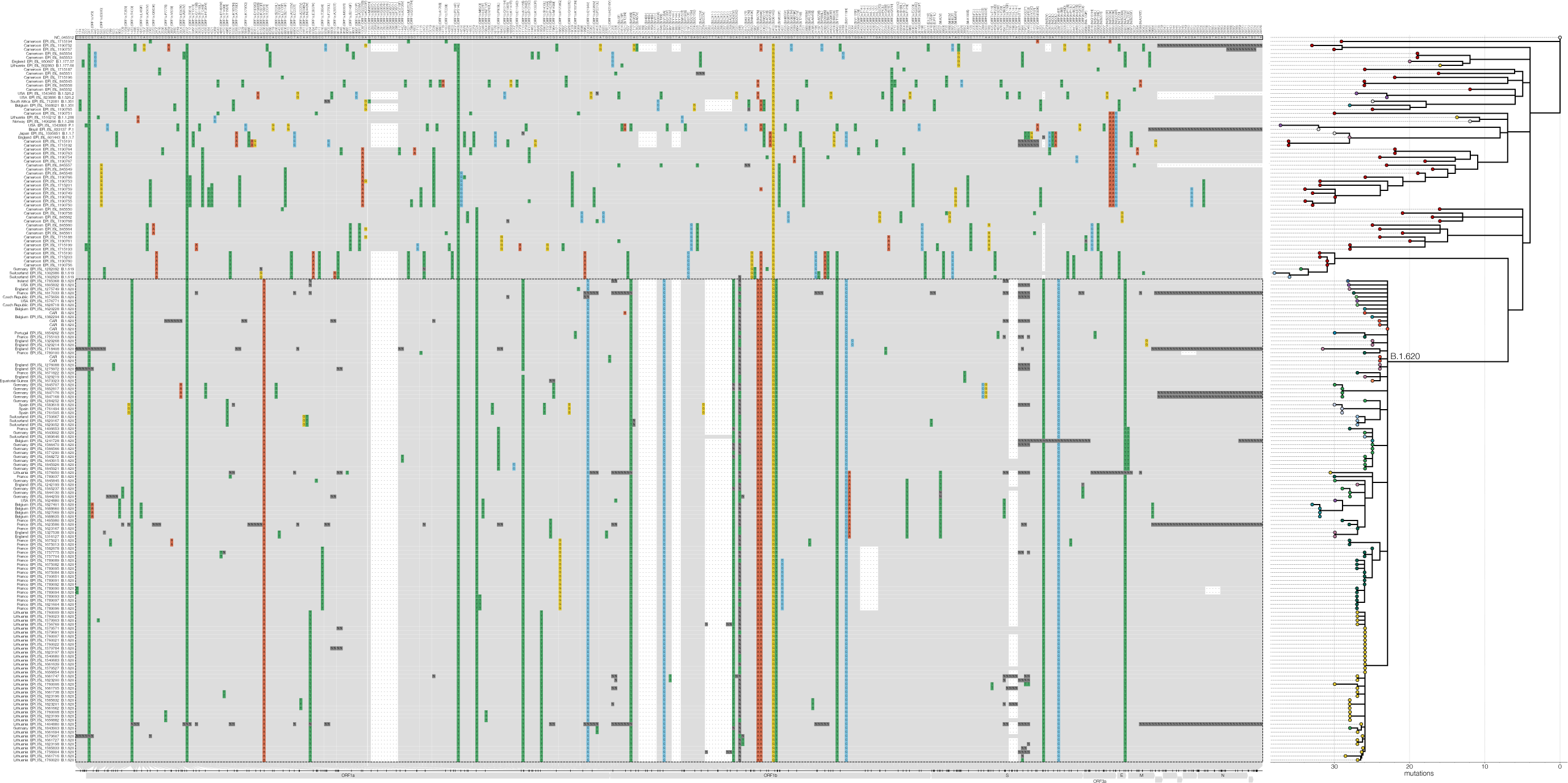
Shared SNPs of investigated lineages. SNP alignment of multiple high-quality genomes from Cameroon, available genomes of lineage B.1.620 as well as earliest and latest genomes of lineages B.1.1.7, B.1.351, P.1, B.1.526.2, B.1.177.60, B.1.177.57, and B.1.1.280. Only SNPs shared by at least two genomes are shown, lineage B.1.620 outlined with a dashed line. Sites identical to the reference (GenBank accession NC 045512) are shown in grey, changes from the reference are indicated and coloured by nucleotide (green for thymidine, red for adenosine, blue for cytosine, yellow for guanine, dark grey for ambiguities, black for gaps). If mutation results in an amino acid change, the column label indicates the gene, reference amino acid, amino acid site, and amino acid change in brackets. Maximum likelihood phylogeny on the right shows the relationships between depicted genomes and was rooted on the reference sequence. Tip circles indicate each genome’s country of origin in the same colour scheme as Figures 3 and 2.

**Figure S5.**
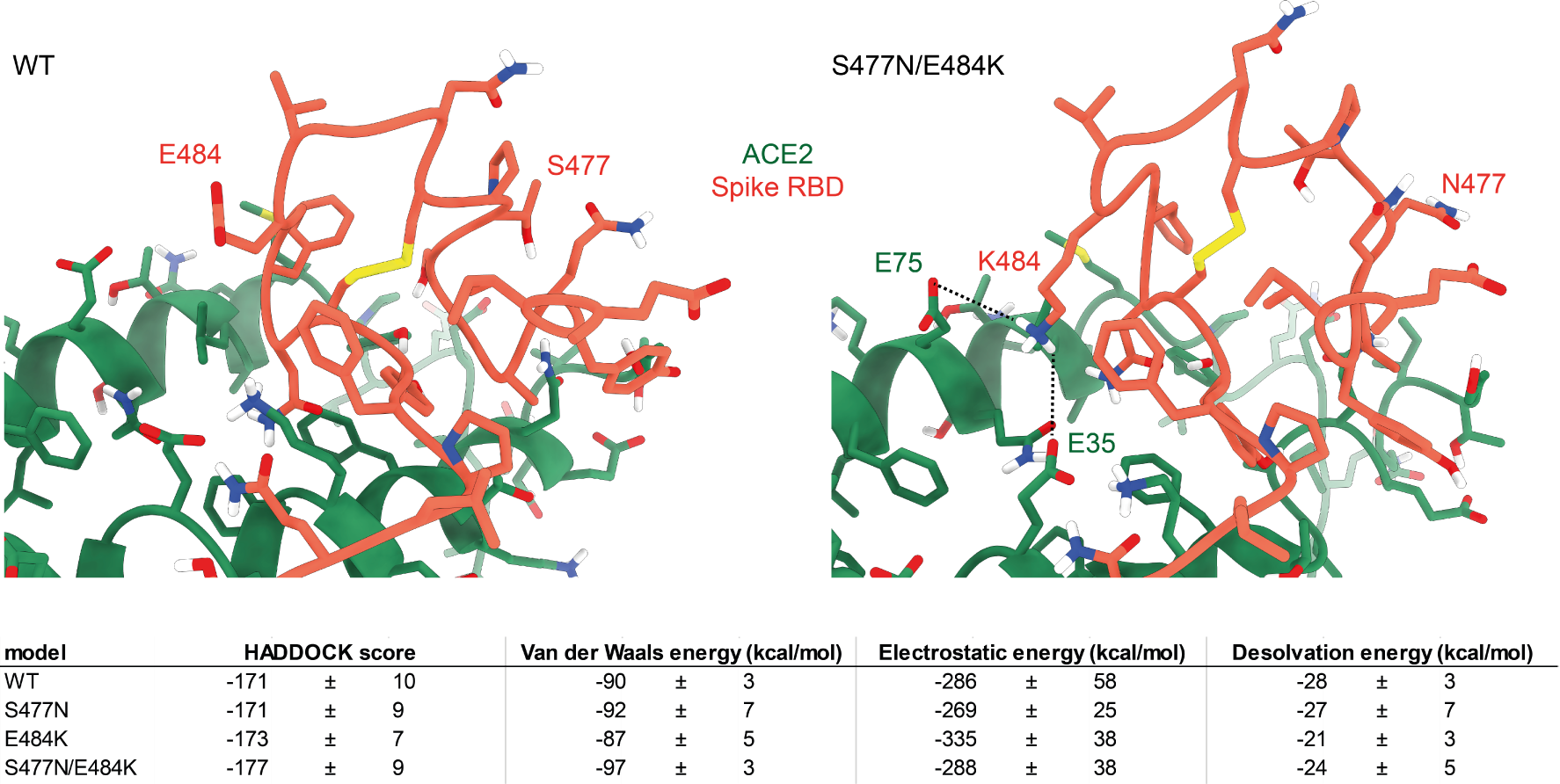
HADDOCK models of ACE2-RBD interaction. HADDOCK scores and energy terms are listed as average +/-standard error of the cluster.

**Figure S6.**
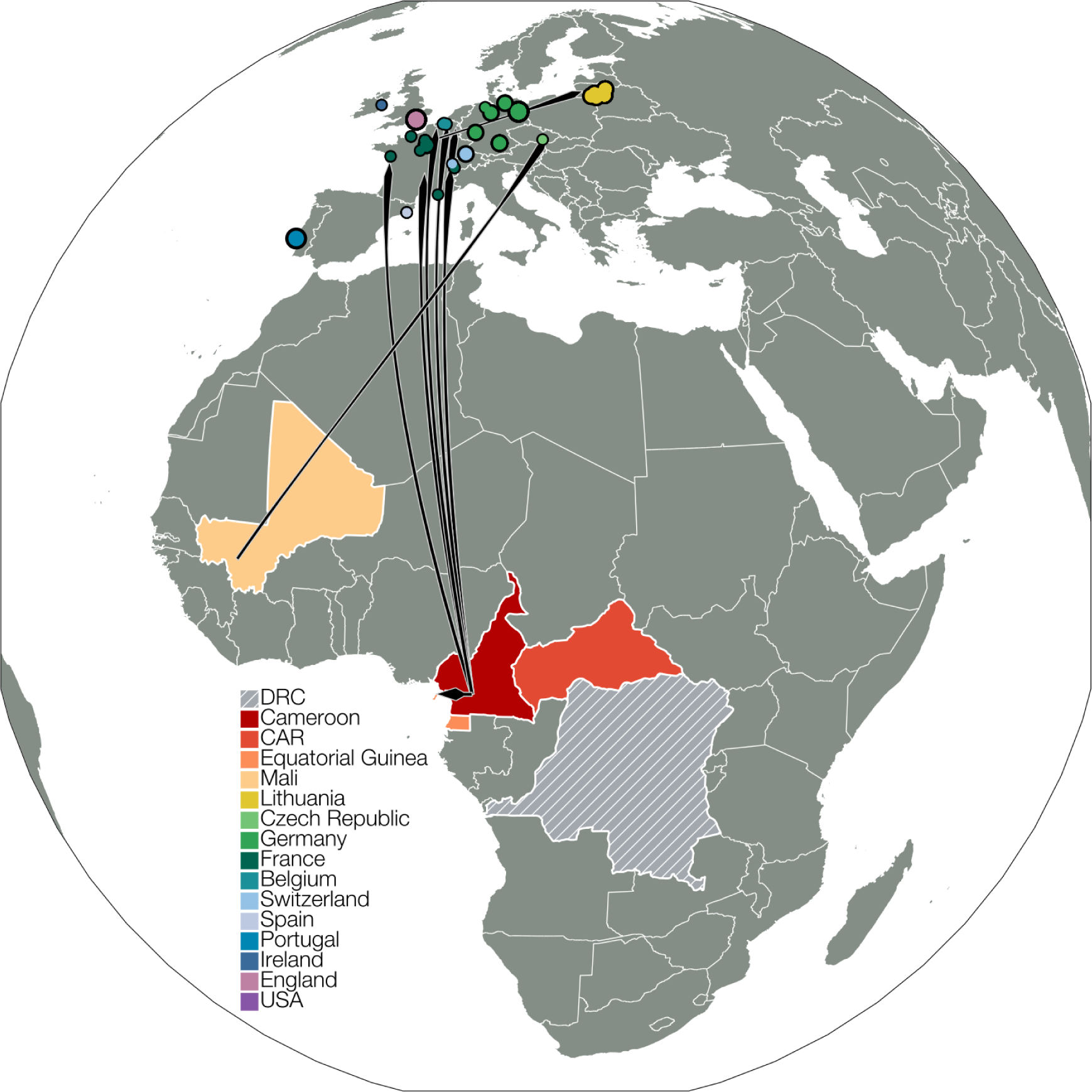
Travel cases and distribution of B.1.620 infections across Africa and Europe. Mali, Cameroon, Central African Republic and Equatorial Guinea are highlighted in Africa and known B.1.620 cases related to travel are displayed as arrows going from known country of origin to destination country. The precise location of B.1.620 cases in Africa is only known for cases from CAR, whose location near the border – near the Dzanga-Sangha Protected Areas – with Cameroon is indicated with a circle. European cases for which no travel information is available and which may represent cases resulting from local transmission are indicated with circles as well. DRC is marked in hatching because B.1.620 from there have been submitted to GISAID, but not used in analyses here.

