## Supplementary material for "Travel-driven emergence and spread of SARS-CoV-2 lineage B.1.620 with multiple VOC-like mutations and deletions in Europe": GISAID acknowledgment table

We gratefully acknowledge the following Authors from the Originating laboratories responsible for obtaining the specimens, as well as the Submitting laboratories where the genome data were generated and shared via GISAID, on which this research is based.

All Submitters of data may be contacted directly via www.gisaid.org

Authors are sorted alphabetically.

| Accession ID | Originating Laboratory | Submitting Laboratory | Authors |
|---|---|---|---|
| EPI_ISL_1008370 | Department of Laboratory Medicine, Division of Clinical Virology, University of Medicine, Vienna | Bergthaler laboratory, CeMM Research Center for Molecular Medicine of the Austrian Academy of Sciences | Lukas Endler, Anna Schedl, Thomas Penz, Benedikt Agerer, Maelle Le Moing, Michael Schuster, Bekir Erguner, Jan Laine, Martin Senekowitsch, Christoph Bock, Andreas Bergthaler |
| EPI_ISL_1008420 | Klinisk mikrobiologi | The Public Health Agency of Sweden | Anna-Malin Linde, Maria Lind Karlberg, Carlo Berg, Oskar Karlsson Lindsjo, Sofia Stamouli, Reza Advani, Mattias Haukland, Petra Holmstrom, Noura Walai, Petra Edquist, Mia Brytting, Anna Risberg, Karin Tegmark-Wisell |
| EPI_ISL_1014570, EPI_ISL_1014617 | Dutch COVID-19 response team | National Institute for Public Health and the Environment (RIVM) | Adam Meijer, Harry Vennema, Dirk Eggink, Jeroen Cremer, Sharon van den Brink, Bas van der Veer, AnneMarie van den Brandt, Florian Zwagemaker, Dennis Schmitz, Chantal Reusken, on behalf of the national COVID-19 response team |
| EPI_ISL_1016855 | Middlemore Hospital | Institute of Environmental Science and Research (ESR) | Xiaoyun Ren, Matt Storey, Nikki Freed, Muhammad Faisal, Jing Wang, Hermes Perez, Anja Werno, Antje van der Linden, Arlo Upton, Chris Mansell, David Hammer, Dragana Drinkovic, Gary McAuliffe, Hana Sofia Andersson, James Ussher, Jill Sherwood, Josh Freeman, Julia Howard, Juliet Elvy, Mary DeAlmeida, Matt Blakiston, Matthew Rogers, Max Bloomfield, Michael Addidle, Michelle Balm, Sally Roberts, Sarah Jefferies, Sharmini Muttaiyah, Susan Morpeth, Susan Taylor, Timothy Blackmore, Vani Sathyendran, Veronica Playle, Virginia Hope, Erasmus Smit, Lauren Jelly, Olin Silander, Joep de Ligt |
| EPI_ISL_1020817 | Laboratory Corporation of America | Respiratory Viruses Branch, Division of Viral Diseases, Centers for Disease Control and Prevention | Peter W. Cook, Dakota Howard, Dhwani Batra, Ben L. Rambo-Martin, Clinton R. Paden, Suxiang Tong, Duncan MacCannell |
| EPI_ISL_1027639 | Department of Microbiology, National Institute for Public Health of Kosova | Charité Universitätsmedizin Berlin, Institut für Virologie | Victor M Corman, Julia Schneider, Donjeta Hajdari, Zana Deva, Xhevat Jakupi, Barbara Mühlemann, Jörn Beheim-Schwarzbach, Talitha Veith, Terry Jones, Christian Drosten |
| EPI_ISL_1038482 | Laboratory Corporation of America | Respiratory Viruses Branch, Division of Viral Diseases, Centers for Disease Control and Prevention | Peter W. Cook, Dakota Howard, Dhwani Batra, Ben L. Rambo-Martin, Clinton R. Paden, Suxiang Tong, Duncan MacCannell |
| EPI_ISL_1055886 | National Laboratory for Health, Environment and Food | CISLD (Clinical Institute of Special Laboratory Diagnostics), University Children's Hospital, University Medical Center Ljubljana | Jernej Kova, Barbara Jenko Bizjan, Tine Tesovnik, Robert Šket, Katarina Kozmos, Ana Grom, Maruša Debeljak, Marko Pokorn, Tadej Battelino |
| EPI_ISL_1061033 | New South Wales Health Pathology Royal Prince Alfred Hospital | Microbiology RPAH | Foster, C.; Au, J.; Ruiz Silva, M.; Deveson, I.; Bull, R.; Van Hal, S.; Rawlinson, W. |
| EPI_ISL_1094328 | MT Public Health Laboratory | Respiratory Viruses Branch, Division of Viral Diseases, Centers for Disease Control and Prevention | Krista Queen, Yan Li, Ying Tao, Jing Zhang, Anna Uehara, Anna Montmayeur, Clinton R. Paden, Peter W. Cook, Rachel Marine, Mili Sheth, Jasmine Padilla, Sarah Nobles, Mark Burroughs, Lori Rowe, Haibin Wang, Ben L. Rambo-Martin, Dhwani Batra, Justin Lee, Suxiang Tong |
| EPI_ISL_1112166 | National Laboratory for Health, Environment and Food, OMM, Maribor | CISLD (Clinical Institute of Special Laboratory Diagnostics), University Children's Hospital, University Medical Center Ljubljana | Jernej Kova, Barbara Jenko Bizjan, Tine Tesovnik, Robert Šket, Katarina Kozmos, Ana Grom, Maruša Debeljak, Marko Pokorn, Tadej Battelino |
| EPI_ISL_1117135 | Instituto Nacional de Saude (INSA) | Instituto Nacional de Saude (INSA) | Borges et al |
| EPI_ISL_1117426 | Nucleo de Pesquisa em Inovacao Terapeutica - UFPE | LABBE, Federal University of Pernambuco | Wilson Jose da Silva Junior, Marcos da Silveira Regueira Neto, Heidi Lacerda Alves da Cruz, Bruno Sampaio, Reginaldo Goncalves de Lima Neto, Maira Galdino da Rocha Pitta, Michelly Cristiny Pereira, Marco Katzenberger, Valdir de Queiroz Balbino |
| EPI_ISL_1120614 | Hospital Margarita Maza de Juarez | CIAD LDM-LGM | Bruno Gome-Gil, Julissa Enciso-Ibarra |
| EPI_ISL_1137621 | Laboratoire central de Virologie | Laboratoire de Biotechnologie | Myriam Seffar, Hakima Kabbaj, Ghizlane EL Amin, Amal Zouaki, Abdelomunim Essabbar, Tarik Aanniz, Mouna Ouadghiri, Saaïd Amzazi, Lahcen Belyamani and Azeddine Ibrahimi |
| EPI_ISL_1161766 | Laboratory Corporation of America | Respiratory Viruses Branch, Division of Viral Diseases, Centers for Disease Control and Prevention | Peter W. Cook, Dakota Howard, Dhwani Batra, Ben L. Rambo-Martin, Minoo Agarwal, Eyad Almasri Debbie Boles, Ayla Burns, Nuthawin Charoensri, Oren Cohen, Susan Countryman, Mary Ann Cristobal, Bobbi Croy, Suzanne Dale, Hrushikesh Deshmukh, Amanda Douglas, Vincent Drouillon, Marcia Eisenberg, Howard Engler, Rama Ghatti, Prashant Gupta, Susan Hicks, Jake Humphrey, Lax Iyer, Manoj Jain, Mohan Kolli, Brian Krueger, Tim Kuphal, Stanley Letovsky, Michael Levandoski, Craig Lukasik, Jonathan Meltzer, Brian Norvell, Mindy Nye, Scott Parker, Christos Petropoulos, John Pruitt, Steven Ragan, Scott Ryan, Mike Sapeta, Jana Schroth, Suresh Babu Selvaraju, Goran Stevovic, Amanda Suchanek, Andrea Throop, Lyndon Tilson, Thomas Urban, Joe Voshell, Kimberly Wagner, Jonathan Williams, Mary Williamson, Qian Zeng, Tricia Zwiefelhofer, Clinton R. Paden, Suxiang Tong, Duncan MacCannell |
| EPI_ISL_1167145 | Iressef Genomics lab | L'institut de Recherche en Santé, de Surveillance Épidémiologique et de Formation (IRESSEF) | Souleymane MBOUP, Abdou PADANE, Abdoulie KANTEH ,Abdul Karim SESAY , Khadim GUEYE, Papa Alassane DIAW, Birahim Piere NDIAYE, Barada CISSE, Aminata MBOUP, Moustapha MBOW, Ndeye Coumba Toure KANE, Nafisatou LEYE, Gora LO, Ambroise AHOUIDI , Astou Gaye GAYE, Aminata DIA, Yacine DIA |
| EPI_ISL_1167764, EPI_ISL_1167777 | Genetica Molecular and Subdepartamento de Virologia ISP Chile | Instituto de Salud Publica de Chile | Javier Tognarelli, Karen Orostica, Barbara Parra, Loredana Arata, Jaime Lagos, Gisselle Barra, Patricia Bustos, Rodrigo Fasce, Andres Castillo, Jorge Fernandez |
| EPI_ISL_1168493, EPI_ISL_1168616 | Instituto de Diagnostico y Referencia Epidemiologicos INDRE_RNLSP | Instituto de Diagnostico y Referencia Epidemiologicos (INDRE) | Claudia Wong-Arambula, Abril Rodriguez-Maldonado, Vanessa Rivero-Arredondo, Ariadna Medina-Benitez, Joaquin Quiroz-Mercado,David Fragoso-Fonseca, Sergio Rangel-Guerrero, Natividad Cruz-Ortiz, Tatiana Nunez-Garcia, Gisela Barrera-Badillo, Lucia Hernandez-Rivas, Irma Lopez-Martinez, Ernesto Ramirez-Gonzalez. |
| EPI_ISL_1181607 | Laboratorio Central de Saude Publica do Estado do Rio de Janeiro (LACEN-RJ) | Laboratory of Respiratory Viruses and Measles, Oswaldo Cruz Institute, FIOCRUZ | Paola Resende, Luciana Appolinario, Fernando Motta, Anna Carolina Paixao, Ana Carolina Mendonca, Alice Sampaio Rocha, Renata Serrano Lopes, Andrea Cony Cavalcanti, Marilda Siqueira on behalf of the Fiocruz COVID-19 Genomic Surveillance Network |
| EPI_ISL_1190749, EPI_ISL_1190750, EPI_ISL_1190751, EPI_ISL_1190752, EPI_ISL_1190753, EPI_ISL_1190754, EPI_ISL_1190755, EPI_ISL_1190756, EPI_ISL_1190757, EPI_ISL_1190758, EPI_ISL_1190759, EPI_ISL_1190760, EPI_ISL_1190761, EPI_ISL_1190762, EPI_ISL_1190763, EPI_ISL_1190764, EPI_ISL_1190765, EPI_ISL_1190766, EPI_ISL_1190767, EPI_ISL_1190768 | | | |
| see above | CREMER(Centre de Rechercherches sur les Maladies Emergentes et Ré-émergentes) | TransVIHMI(Recherches Translationnelles sur le VIH et les Maladies Infectieuses) | Celestin Godwe, Christelle Butel, Martin Maidadi Foudi, Laetitia Serrano, Dowbiss Meta Djomsi, Nicole Vidal, Esemu Livo, Rodrigue Kamga, Marie Amougou , Eitel Mpoudi Ngole,Ahidjo Ayouba, Marcel Tongo, Martine Peeters, Eric Delaporte |
| EPI_ISL_1195207 | National Public Health Center, COVID Laboratory | National Public Health Center, National Biosafety Laboratory | Bernadett Pályi, Zoltán Kis, Nóra Magyar, Judit Henczkó, Dániel Déri, Norbert Solymosi |
| EPI_ISL_1208402 | ACT Pathology | Schwessinger Lab | Ashley Jones, Benjamin Schwessinger, Robert Lanfear, Megan McDonald, Ming-Dao Chia, Kevin Murray, Robyn N Hall, Craig Kennedy, Karina Kennedy |
| EPI_ISL_1214521 | SYNLAB MVZ Leverkusen | Robert Koch Institute | unknown |
| EPI_ISL_1216076 | MRCG at LSHTM Genomics lab | MRCG at LSHTM Genomics lab | Abdul Karim sesay, Abdoulie Kanteh, Jarra Manneh, Mariama Kujabi, Bakary Sanyang |
| EPI_ISL_1233103 | Centre for Dengue Research and AICBU, Department of Immunology and Molecular Medicine | Centre for Dengue Research and AICBU, Department of Immunology and Molecular Medicine | Chandima Jeewandara, Deshni Jayathilaka, Dinuka Ariyaratne, Tibutius Thanesh Pramanayagam, Diyanath Ranasinghe, Laksiri Gomes, Gathsaurie Neelika Malavige |
| EPI_ISL_1239005 | GA Department of Public Health | GA Department of Public Health | Stacy Reeves, Jonathan Edwards, Cynthia Dixey, Tonia Parrott, Aliyah Fields, Taylor Smith |
| EPI_ISL_1241728 | SYNLAB | GIGA Medical Genomics | Keith Durkin, Maria Artesi, Sébastien Bontems, Raphaël Boreux, Bouchra Boujemla, Nathalie Renotte, Cécile Meex, Pierrette Melin, Marie-Pierre Hayette, |

| | | | |
|---|---|---|---|
| | | | Vincent Bours |
| EPI_ISL_1242199 | Lighthouse Lab in Cambridge | Wellcome Sanger Institute for the COVID-19 Genomics UK (COG-UK) Consortium | Rob Howes, The Lighthouse Lab in Cambridge and Alex Alderton, Roberto Amato, Jeffrey Barrett, Sonia Goncalves, Ewan Harrison, David K. Jackson, Ian Johnston, Dominic Kwiatkowski, Cordelia Langford, John Sillitoe on behalf of the Wellcome Sanger Institute COVID-19 Surveillance Team |
| EPI_ISL_1253511 | Quest Diagnostics Incorporated | Centers for Disease Control and Prevention Division of Viral Diseases, Pathogen Discovery | Peter W. Cook, Dakota Howard, Dhwani Batra, Ben L. Rambo-Martin, S. H. Rosenthal, A. Gerasimova, R. M. Kagan, B. Anderson, M. Hua, Y. Liu, L.E. Bernstein, K.E. Livingston, A. Perez, I. A. Shlyakhter, R. V. Rolando, R. Owen, P. Tanpaiboon, F. Lacbawan, Clinton R. Paden, Suxiang Tong, Duncan MacCannell |
| EPI_ISL_1255144, EPI_ISL_1255271 | West African Centre for Cell Biology of Infectious Pathogens (WACCBIP), University of Ghana, Accra, Ghana | West African Centre for Cell Biology of Infectious Pathogens (WACCBIP), University of Ghana, Volta Road, Legon-Accra, Ghana | Collins M. Morang'a, Joyce M. Ngoi, Evelyn B. Quansah, Samirah Saiid, Dominic S.Y. Amuzu, Vincent Appiah, Philip M. Soglo, Vanessa Magnussen, Aisha Mohammed, Kesego Tapela, Nelson Kibinge, Abdoulaye B Diallo, Frederick Kumi-Ansah, Theophilus Odoom, Oliver D Boakye5, Emmanuella Amoako4, Abdul-Karim Abass, , Samuel Kaba Akoriyea, Frederick Tei-Maya, Lucas N. Amenga-Etego, Dam Kenneth Mibut, Yaw Bediako, Benjamin Demah Nuertey, Gordon A Awandare, Peter K Quashie, Gordon A Awandare, Yaw Bediako |
| EPI_ISL_1261374 | National Institute of Public Health | National Reference Laboratory for Influenza and Respiratory Viruses CZE | Helena Jirincova, Jaromira Vecerova, Timotej Suri, Dusan Trnka, Alexander Nagy |
| EPI_ISL_1265458 | BTKLPP Kelas I Makassar | Eijkman Institute for Molecular Biology, Ministry of Research and Technology/National Agency for Research and Innovation; National Institute of Health Research and Development | Sukma Oktavianthi, Lydia V. Panggalo, Edison Johar, Hidayat Trimarsanto, Frilasita A Yudhaputri, Iskandar Adnan, Willy Agustine, Slamet, Vivi Setiawaty, Hana Apsari Pawestri, Safarina G Malik, Khin Saw Myint, Amin Soebandrio |
| EPI_ISL_1273393 | National Reference Laboratory - Ministry of Health Maseru Lesotho | National Institute for Communicable Diseases of the National Health Laboratory Service | Gorova V, Mathabo M, Mooko M, Banda R, Amoako DG, Scheepers C, Mohale T, Ntuli N, Mahlangu B, Ismail A, Bhiman JN |
| EPI_ISL_1275749, EPI_ISL_1275972, EPI_ISL_1276088 | Lighthouse Lab in Cambridge | Wellcome Sanger Institute for the COVID-19 Genomics UK (COG-UK) Consortium | Rob Howes, The Lighthouse Lab in Cambridge and Alex Alderton, Roberto Amato, Jeffrey Barrett, Sonia Goncalves, Ewan Harrison, David K. Jackson, Ian Johnston, Dominic Kwiatkowski, Cordelia Langford, John Sillitoe on behalf of the Wellcome Sanger Institute COVID-19 Surveillance Team |
| EPI_ISL_1278684 | Clalit Health Services Laboratories, Israel | Stern Lab | Stern Lab |
| EPI_ISL_1282092 | Bioscientia Labor Wermsdorf | Robert Koch Institute | unknown |
| EPI_ISL_1284252 | SYNLAB MVZ Trier | Robert Koch Institute | unknown |
| EPI_ISL_1288215, EPI_ISL_1288383 | Laboratorio Central de Epidemiología (LCE) | Instituto de Biotecnología de la UNAM | Consorcio Mexicano de Vigilancia Genómica (CoViGen-Mex). Authors (in alphabetical order): Julio Elias Alvarado-Yaah, Carlos F. Arias, Santiago Ávila-Ríos, Víctor Hugo Borja-Aburto, Celia Boukadida, Juan Bautista Chale-Dzul , José Antonio Enciso-Moreno, Gloria Elena Espinoza-Ayala, Fernando Fontove-Herrera, Concepción Grajales-Muñiz, Ricardo Grande, Alfredo Herrera-Estrella, Carla Ivón Herrera-Najera, Pavel Isa, Brenda Irasema Maldonado-Meza, Bernardo Martínez-Miguel, Margarita Matías-Florentino, María Guadalupe de Jesús Mireles-Rivera, Gloria María Molina-Salinas, Hector Montoya-Fuentes, José Esteban Muñoz-Medina, José de Jesús Nuñez-Contreras, Alicia Ocaña-Mondragón, Luis Alberto Ochoa-Carrera, Hector Esteban Paz-Juárez, Francisco Pulido, Helen Haydee Fernanda Ramírez-Plascencia, Angel Gustavo Salas-Lais, Jorge Ivan Salinal-Nevarez, Alejandro Sanchez-Flores, Clara Esperanza Santacruz-Tinoco, María Guadalupe Santiago-Mauricio , Nelly Sélem-Mojica, Blanca Taboada , Gloria Vazquez |
| EPI_ISL_1292663 | State Laboratories Division, Hawaii State Department of Health | State Laboratories Division, Hawaii State Department of Health | Pamela O'Brien, Drew Kuwazaki, Ayana Garnet, Razvan Sultana, Edward Desmond |
| EPI_ISL_1300526 | Public Health Virology-Forensic and Scientific Services (PHV-FSS) | Public Health Virology-Forensic and Scientific Services (PHV-FSS) | Son Nguyen |
| EPI_ISL_1302341 | Laboratorio Central de Epidemiología IMSS | Instituto de Biotecnología de la UNAM | Authors from IBT, IMSS, InDRE and INER (in alphabetical order): Carlos F. Arias, Santiago Ávila-Ríos, Gisela Barrera-Badillo, Eduardo Becerril-Vargas, Celia Boukadida, Natividad Cruz-Ortiz, Larissa Fernandes-Matano, Ricardo Grande, Lucia Hernandez-Rivas, Alejandra Hernández-Terán, Pavel Isa, Irma Lopez-Martinez, José Arturo Martínez-Orozco, Margarita Matías-Florentino, Fidencio Mejía-Nepomuceno, Edgar Mendieta-Condado, Mario Mújica-Sánchez, José Esteban Muñoz-Medina, Tatiana Nunez-Garcia, Luis Alberto Ochoa-Carrera, Hector Esteban Paz-Juárez, Francisco Pulido, José Ernesto Ramírez-González, Alma Rincón-Rubio, Teresita Rojas-Mendoza, Jorge Salas-Hernández, Alejandro Sanchez-Flores, Clara Esperanza Santacruz-Tinoco, Andrea Santos Coy-Arechavaleta, Blanca Taboada, Gloria Vazquez, Joel Armando Vázquez-Pérez, Jerome Jean Verleyen |
| EPI_ISL_1302680 | Nucleic Acid Testing, National Reference Laboratory | GIGA Medical Genomics | Yvan Butera, Keith Durkin, Maria Artesi, Bouchra Boujemla, Robert Rutayisire, Patrick Tuyisenge, Esperence Umumararungu, Sébastien Bontems, Marie-Pierre Hayette, Nathalie Renotte, Swaibu Gatare, Jacob Souopgui, Sabin Nsanzimana, Vincent Bours, Léon Mutesa |
| EPI_ISL_1302829, EPI_ISL_1302986 | ADMED Microbiologie | Genomics and Transcriptomics, Philip Morris International | Reto Lienhard, Marie-Lise Tritten, Emmanuel Guedj, Nicolas Sierro, Rémi Dulize, David Bornand, Mehdi Auberson, Maxime Berthouzoz, Nikolai Ivanov, Manuel Peitsch |
| EPI_ISL_1303367 | Instituto Nacional de Salud- Dirección de Redes de Laboratorios de Salud Pública | Instituto Nacional de Salud- Dirección de Investigación en Salud Pública | Katherine Laiton-Donato, Diego A. Álvarez-Díaz, Carlos Franco-Muñoz, Hector Alejandro Ruiz-Moreno, Maria T. Herrera-Sepúlveda, Diego Andrés Prada, Jhonnatan Reales-González, Sheryll Corchuelo, Julian Naizaque, Gerardo Santamaría, Sergio Gomez, Lisseth Pardo, Juan Camilo Martinez, Marta Lopez Blanco, Ángela Alarcon Cruz, Diana Malo, Carmen Osorio, Magdalena Wiesner, Martha Lucia Ospina Martinez, Marcela Mercado-Reyes |
| EPI_ISL_1314567 | Quest Diagnostics Incorporated | Centers for Disease Control and Prevention Division of Viral Diseases, Pathogen Discovery | Peter W. Cook, Dakota Howard, Dhwani Batra, Ben L. Rambo-Martin, S. H. Rosenthal, A. Gerasimova, R. M. Kagan, B. Anderson, M. Hua, Y. Liu, L.E. Bernstein, K.E. Livingston, A. Perez, I. A. Shlyakhter, R. V. Rolando, R. Owen, P. Tanpaiboon, F. Lacbawan, Clinton R. Paden, Suxiang Tong, Duncan MacCannell |
| EPI_ISL_1315311 | Middlemore Hospital | Institute of Environmental Science and Research (ESR) | Rachel Boyle, SallyAnn Harbison, Olivia Stroeven, Xiaoyun Ren, Matt Storey, Nikki Freed, Muhammad Faisal, Jing Wang, Hermes Perez, Anja Werno, Antje van der Linden, Arlo Upton, Chris Mansell, David Hammer, Dragana Drinkovic, Gary McAuliffe, Hana Sofia Andersson, James Ussher, Jill Sherwood, Josh Freeman, Julia Howard, Juliet Elvy, Mary DeAlmeida, Matt Blakiston, Matthew Rogers, Max Bloomfield, Michael Addidle, Michelle Balm, Sally Roberts, Sarah Jefferies, Sharmini Muttaiyah, Susan Morpeth, Susan Taylor, Timothy Blackmore, Vani Sathyendran, Veronica Playle, Virginia Hope, Erasmus Smit, Lauren Jelly, Olin Silander, Joep de Ligt |
| EPI_ISL_1316127 | Lighthouse Lab in Cambridge | Wellcome Sanger Institute for the COVID-19 Genomics UK (COG-UK) Consortium | Rob Howes, The Lighthouse Lab in Cambridge and Alex Alderton, Roberto Amato, Jeffrey Barrett, Sonia Goncalves, Ewan Harrison, David K. Jackson, Ian Johnston, Dominic Kwiatkowski, Cordelia Langford, John Sillitoe on behalf of the Wellcome Sanger Institute COVID-19 Surveillance Team |
| EPI_ISL_1318472 | MT Public Health Laboratory | Centers for Disease Control and Prevention Division of Viral Diseases, Pathogen Discovery | Krista Queen, Yan Li, Ying Tao, Jing Zhang, Anna Uehara, Anna Montmayeur, Clinton R. Paden, Kristen Knipe, Matthew Schmerer, Shoshona Le, Katie Dillon, Peter W. Cook, Rachel Marine, Mili Sheth, Jasmine Padilla, Sarah Nobles, Mark Burroughs, Lori Rowe, Haibin Wang, Ben L. Rambo-Martin, Kristine Lacek, Sam Shepard, Dhwani Batra, Suxiang Tong, Justin Lee |
| EPI_ISL_1327538, EPI_ISL_1329214, EPI_ISL_1329219, EPI_ISL_1329268 | Lighthouse Lab in Cambridge | Wellcome Sanger Institute for the COVID-19 Genomics UK (COG-UK) Consortium | Rob Howes, The Lighthouse Lab in Cambridge and Alex Alderton, Roberto Amato, Jeffrey Barrett, Sonia Goncalves, Ewan Harrison, David K. Jackson, Ian Johnston, Dominic Kwiatkowski, Cordelia Langford, John Sillitoe on behalf of the Wellcome Sanger Institute COVID-19 Surveillance Team |
| EPI_ISL_1363477 | Sonora Quest Laboratories | TGen North | Jolene Bowers, Heather Centner, Chris French, Hayley Yaglom, Ashlyn Pfeiffer, Darrin Lemmer, Dave Engelthaler, The Arizona COVID Genomics Union (ACGU) |
| EPI_ISL_1365031 | Molecular diagnostic unit for viral haemorrhagic fevers and emerging viruses, Bouaké CHU Laboratory | Molecular diagnostic unit for viral haemorrhagic fevers and emerging viruses, Bouaké CHU Laboratory | Chantal Akoua-Koffi, Diané Bamourou, Etilé Anoh, Oby Wayoro, Safiatou Karidioula, Adjaratou Traoré, Soundélé Maïté, Monemo Pacome, Coulibaly Mbegnan, Bamba Fatoumata Touré, Kra Ouffoué, Grit Schubert, Essia Belarbi, Fabian Leendertz |
| EPI_ISL_1366745 | OUCRU | OUCRU | Nguyen Van Vinh Chau, Nguyen Thi Thu Hong, Nghiem My Ngoc, Nguyen To Anh, Huynh Trung Trieu, Le Nguyen Truc Nhu, Lam Minh Yen, Ngo Ngoc Quang Minh, Nguyen Thanh Phong, Nguyen Thanh Truong, Le Thi Thu Huong, Tran Nguyen Hoang Tu, Le Manh Hung, Tran Tan Thanh, Nguyen Thanh Dung, Nguyen Tri Dung, Guy Thwaites, Le Van Tan |
| EPI_ISL_1367678, EPI_ISL_1367685 | Molecular diagnostic unit for viral haemorrhagic fevers and emerging viruses, Bouaké CHU Laboratory | Molecular diagnostic unit for viral haemorrhagic fevers and emerging viruses, Bouaké CHU Laboratory | Chantal Akoua-Koffi, Diané Bamourou, Etilé Anoh, Oby Wayoro, Safiatou Karidioula, Adjaratou Traoré, Soundélé Maïté, Monemo Pacome, Coulibaly Mbegnan, Bamba Fatoumata Touré, Kra Ouffoué, Grit Schubert, Essia Belarbi, Fabian Leendertz |
| EPI_ISL_1369646 | University Hospitals of Geneva, Laboratory of Virology | HUG, Laboratory of Virology and the Health2030 Genome Center | Samuel Cordey, Ana Rita Goncalves, Laurent Kaiser, Lorenzo Cerutti, Henri Pegeot, Melyssa Elies, Deborah Penet, Keith Harshman, Ioannis Xenarios, Emmanouil Dermitzakis |

| | | | |
|---|---|---|---|
| EPI_ISL_1371900 | C H DE LA POLYNESIE FRANCAISE | CNR Virus des Infections Respiratoires - France SUD | Antonin Bal, Gregory Destras, Gwendolyne Burfin, Hadrien Regue, Quentin Semanas, Martine Valette, Bruno Lina, Laurence Josset |
| EPI_ISL_1381065 | IAL Regional de Santo Andre | Instituto Adolfo Lutz, Interdiciplinary Procedures Center, Strategic Laboratory | Claudio Tavares Sacchi, Claudia Regina Gonçalves, Erica Valessa Ramos Gomes, Karoline Rodrigues Campos, Caio Vinicius Dias Lopes |
| EPI_ISL_1381254 | Centro de Investigacion Biomedica del Noreste (CIBIN) | Unidad de Genomica Avanzada | Consorcio Mexicano de Vigilancia Genomica (CoViGen-Mex). Authors (in alphabetical order): Julio Elias Alvarado-Yaah, Carlos F. Arias, Santiago Avila-Rios, Victor Hugo Borja-Aburto, Celia Boukadida, Juan Bautista Chale-Dzul , Jose Antonio Enciso-Moreno, Gloria Elena Espinoza-Ayala, Fernando Fontove-Herrera, Concepcion Grajales-Muniz, Ricardo Grande, Alfredo Herrera-Estrella, Carla Ivon Herrera-Najera, Pavel Isa, Brenda Irasema Maldonado-Meza, Bernardo Martinez-Miguel, Margarita Matias-Florentino, Maria Guadalupe de Jesus Mireles-Rivera, Gloria Maria Molina-Salinas, Hector Montoya-Fuentes, Jose Esteban Munoz-Medina, Jose de Jesus Nunez-Contreras, Alicia Ocana-Mondragon, Luis Alberto Ochoa-Carrera, Hector Esteban Paz-Juarez, Francisco Pulido, Helen Haydee Fernanda Ramirez-Plascencia, Angel Gustavo Salas-Lais, Jorge Ivan Salinal-Nevarez, Alejandro Sanchez-Flores, Clara Esperanza Santacruz-Tinoco, Maria Guadalupe Santiago-Mauricio, Nelly Selem-Mojica, Blanca Taboada, Gloria Vazquez |
| EPI_ISL_1382294 | KU Leuven, Rega Institute, Clinical and Epidemiological Virology | KU Leuven, Rega Institute, Clinical and Epidemiological Virology | Tony Wawina-Bokalanga, Bert Vanmechelen, Joan Marti-Carerras, Piet Maes |
| EPI_ISL_1385807 | Alfa Diagnostica LLC | ONCOGENE LLC | ONCOGENE LLC |
| EPI_ISL_1396345 | Laboratorio del Hospital Regional Ushuaia Gdor. Ernesto Campos | Nodo de Secuenciación Tierra del Fuego - Hospital Regional Ushuaia - Centro Austral De Investigaciones Científicas - Universidad Nacional De Tierra Del Fuego on behalf of 'Proyecto Argentino Interinstitucional de genomica de SARS-CoV-2' (PAIS Consortium) | Carina Andrea De Roccis, Gabriel Alejandro Castro, Silvana Beatriz Cáceres, Carolina Beatriz Yulan, Manuel Fabian Boutureira, Alejandro Ezequiel Rojas, Fernando Gallego, Santiago Guillermo Ceballos, Cristina Fernanda Nardi, Ivan Dario Gramundi |
| EPI_ISL_1400539 | WHO National Influenza Centre Russian Federation | WHO National Influenza Centre Russian Federation | Andrey Komissarov, Artem Fadeev, Anna Ivanova, Kseniya Komissarova, Dmitry Bazhenov, Mikhail Bakaev, Daria Danilenko, Ksenia Safina, Elena Nabieva, Georgii Bazykin, Dmitry Lioznov |
| EPI_ISL_1404880 | National Institute for Food and Veterinary Risk Assessment | Lithuanian University of Health Sciences | Arnoldas Pautienius, Kamile Tamusauskaite, Gediminas Alzbutas, Dovydas Gecys, Lukas Zemaitis, Vaiva Lesauskaite |
| EPI_ISL_1406391 | Department of Virology | Department of Virology | Massab Umair, Aamer Ikram, Muhammad Salman, Nazish Badar, Sana Tamim, Zaira Rehman, Abdul Ahad, Shannon Whitmer, Melissa Mobley, Austin Leach, Ketan Patel, Joel Montgomery, John Klena |
| EPI_ISL_1406653 | Centre Hospitalier Universitaire Clermont-Ferrand | CHU Clermont-Ferrand, service de virologie | Bisseux Maxime, Mirand Audrey, Combes Patricia, Henquell Cécile |
| EPI_ISL_1407196 | National HIV Reference Laboratory, Ministry of Health, Public Health Institute of Malawi | KRISP, KZN Research Innovation and Sequencing Platform | Mvula B, Chilima B, Chiwaula M, Mwangomba W, Panja L, Kasambara W, Auld A, Kim L, Kampira E, Kaba M, Wadonda N, Maida A, Giandhari J, Pillay S, Naidoo Y, Lessells R, Emmanuel SJ, Tegally H, Wilkinson E, de Oliveira T |
| EPI_ISL_1415344 | National Centre For Cell Science | National Centre For Cell Science - INSACOG | Dhiraj Paul, Mitali Inamdar, Sonal Manik Chavan, Mohak P Gujare, Shivang P. Bhanushali, Manoj Kumar Bhat, Ajay Pillai, INSACOG Consortium team, Yogesh Shouche. |
| EPI_ISL_1416681 | Laboratorio Central de Epidemiología (LCE) | Instituto de Biotecnología de la UNAM | Consorcio Mexicano de Vigilancia Genómica (CoViGen-Mex). Authors (in alphabetical order): Julio Elias Alvarado-Yaah, Carlos F. Arias, Santiago Ávila-Ríos, Víctor Hugo Borja-Aburto, Celia Boukadida, Juan Bautista Chale-Dzul , José Antonio Enciso-Moreno, Gloria Elena Espinoza-Ayala, Fernando Fontove-Herrera, Concepción Grajales-Muñiz, Ricardo Grande, Alfredo Herrera-Estrella, Carla Ivón Herrera-Najera, Pavel Isa, Brenda Irasema Maldonado-Meza, Bernardo Martínez-Miguel, Margarita Matías-Florentino, María Guadalupe de Jesús Mireles-Rivera, Gloria María Molina-Salinas, Hector Montoya-Fuentes, José Esteban Muñoz-Medina, José de Jesús Nuñez-Contreras, Alicia Ocaña-Mondragón, Luis Alberto Ochoa-Carrera, Hector Esteban Paz-Juárez, Francisco Pulido, Helen Haydee Fernanda Ramírez-Plascencia, Angel Gustavo Salas-Lais, Jorge Ivan Salinal-Nevarez, Alejandro Sanchez-Flores, Clara Esperanza Santacruz-Tinoco, María Guadalupe Santiago-Mauricio , Nelly Sélem-Mojica, Blanca Taboada , Gloria Vazquez |
| EPI_ISL_1417511 | Swedish national genomic surveillance program of SARS-CoV-2 | The Public Health Agency of Sweden | Swedish national genomic surveillance program of SARS-CoV-2 |
| EPI_ISL_1419722 | INSACOG-WB | National Institute of Biomedical Genomics - INSACOG | Arindam Maitra, Bhaswati Bandyopadhyay, Nidhan Kumar Biswas, Tamal Ghosh, Sreedhar Chinnaswamy, Ajay Chakraborti, Saumitra Das |
| EPI_ISL_1424529, EPI_ISL_1424599 | Queensland Medical Laboratories | Victorian Infectious Diseases Reference Laboratory (VIDRL) and the Melbourne Diagnostic Unit Public Health Laboratory (MDU-PHL) | Palou, T., Vaccher, S., Seemann, T., Sherry, N.L. |
| EPI_ISL_1440264 | KEMRI-Wellcome Trust Research Programme,Kilifi | KEMRI-Wellcome Trust Research Programme,Kilifi | Githinji G.,Mohamed K.S.,deLaurent Z.,Mburu M.W. |
| EPI_ISL_1443661 | Institute of Microbiology, Universidad San Francisco de Quito | Omics Sciences Laboratory | Derly Andrade Molina, Rubén Armas González, Gabriel Morey León, Darlyn Amaya, Katheryn Sacheri Viteri, Emily Sulay Saltos Montalvo, Paula Juliana Gavilanes Jarrín, Sully Márquez,, Fernanda Zurita, Juan José Guadalupe, Monica Becerra-Wong, Belén Prado-Vivar, Bernardo Gutiérrez, Andrea Cunguan, Nabih Dahik, Dayron Brossad, Patricio Rojas-Silva, Gabriel Trueba, Michelle Grunauer, Verónica Barragán, Paúl Cárdenas, Juan Carlos Fernández Cadena |
| EPI_ISL_1446920 | NC State Laboratory of Public Health | Centers for Disease Control and Prevention Division of Viral Diseases, Pathogen Discovery | Mili Sheth, Sarah Nobles, Jasmine Padilla, Mark Burroughs, Shoshona Le, Katie Dillon, Peter Cook, Clinton R. Paden, Dhwani Batra, Krista Queen, Kristen Knipe, Dakota Howard, Yvette Unoarumhi, Darlene Wagner, Matthew Schmerer, Ben L. Rambo-Martin, Kristine Lacek, Sam Shepard, Alison Laufer Halpin, Dave Wentworth, Vivien Dugan, Suxiang Tong, Justin Lee |
| EPI_ISL_1456512 | Dutch COVID-19 response team | National Institute for Public Health and the Environment (RIVM) | Adam Meijer, Harry Vennema, Dirk Eggink, Jeroen Cremer, Sharon van den Brink, Bas van der Veer, AnneMarie van den Brandt, Lisa Wijsman, Kim Freriks, Ryanne Jaarsma, Euníce Then, Jolienke Hardeman, Lynn Aarts, Sanne Bos, Melissa van Tuil, Robert Kohl, Linda van de Nes, Sjoerd Kuiling, James Groot, Florian Zwagemaker, Dennis Schmitz, Annelies Kroneman, Karim Hajji, Chantal Reusken, on behalf of the national COVID-19 response team |
| EPI_ISL_1461009 | CNR Virus des Infections Respiratoires - France SUD | CNR Virus des Infections Respiratoires - France SUD | Antonin Bal, Gregory Destras, Gwendolyne Burfin, Hadrien Regue, Quentin Semanas, Martine Valette, Bruno Lina, Laurence Josset |
| EPI_ISL_1468846 | Sharp HealthCare Laboratory | Andersen lab at Scripps Research | SEARCH Alliance San Diego with Aaron Harding, Jacquelyn Berumen, Cathy Woerle, Liam McGinnis, Art Mendoza, Omid Bakhtar |
| EPI_ISL_1469358, EPI_ISL_1469380 | MRC/UVRI & LSHTM Uganda Research Unit | Where sequence data have been generated and submitted to GISAID | Matthew Cotten, Dan Lule Bugembe, My V.T. Phan, Isaac Sseeewanyana, Patrick Semanda, Susan Nabadda, Pontiano Kaleebu |
| EPI_ISL_1482701 | MUSC Molecular Pathology Laboratory | MUSC Molecular Pathology Laboratory | Julie W. Hirschhorn, W. Bailey Glen Jr, Dariusz Pytel, Jaclyn Dunne, Kristen Maurer, Frederick S. Nolte |
| EPI_ISL_1483032 | LESP Nuevo Leon | Instituto de Diagnostico y Referencia Epidemiologicos (INDRE) | Claudia Wong-Arambula, Abril Rodriguez-Maldonado, Vanessa Rivero-Arredondo, Ariadna Medina-Benitez, Joaquin Quiroz-Mercado, Sergio Rangel-Guerrero, Natividad Cruz-Ortiz, Tatiana Nunez-Garcia, Gisela Barrera-Badillo, Lucia Hernandez-Rivas, Irma Lopez-Martinez, Ernesto Ramirez-Gonzalez. |
| EPI_ISL_1490266 | Ostfold Hospital Trust - Kalnes, Centre for Laboratory Medicine, Section for gene technology and infection serology | Norwegian Institute of Public Health, Department of Virology | Kathrine Stene-Johansen, Kamilla Heddeland Instefjord, Hilde Elshaug, Garcia Llorente Ignacio, Jon Bråte, Engebretsen Serina Beate,Pedersen Benedikte Nevjen, Debech Nadia, Atiya R Ali,Marie Paulsen Madsen, Rasmus Riis Kopperud, Hilde Vollan, Karoline Bragstad, Olav Hungnes |
| EPI_ISL_1490697 | Unity Health Toronto | Ontario Institute for Cancer Research | Ramzi Fattouh, Larissa M. Matukas, Yan Chen,Mark Downing, Trina Otterman, Karel Boissinot, Le Luu, Samira Mubareka, TIBDN, Ilinca Lungu, Bernard Lam, Jeremy Johns, Paul Krzyzanowski, Richard de Borja, Felicia Vincelli, Philip Zuzarte, Jared T. Simpson |
| EPI_ISL_1495980 | Laboratoire Biorylis | National Reference Center for Viruses of Respiratory Infections, Institut Pasteur, Paris | Marion Barbet, Sylvie Behillil, Méline Bizard, Angela Brisebarre, Camille Capel, Louise Lefrançois, Etienne Simon-Lorière, Vincent Enouf, Maud Vanpeene, Sylvie van der Werf, Potiron Grégoire |
| EPI_ISL_1498716 | BioneXt Lab | Laboratoire national de sante, Microbiology, Microbial Genomics Platform | Anke Wienecke-Baldacchino, Catherine Ragimbeau,Jessica Tapp, Fatu Djabi, Lise Pignon, Raoul Salmon, Thibault Ferrandon, Tamir Abdelrahman |
| EPI_ISL_1502178 | Division of Medical Virology, National Health Laboratory Service (NHLS), Tygerberg Hospital / Stellenbosch University | Division of Medical Virology, Stellenbosch University and NHLS Tygerberg Hospital | Susan Engelbrecht, Bronwyn Kleinhans, Gert van Zyl, Wolfgang Preiser |
| EPI_ISL_1502988 | Gorgas Memorial Laboratory of Health Studies | Gorgas Memorial Laboratory of Health Studies | Gonzalez Claudia, Leyda Abrego, Moreno Ambar, Oris Chavarria, Jessica Gondola, Marlenne Castillo, Ortiz Alma, Castillo Jorge, Moreno Brechla, Franco Danilo, Lopez-Verges Sandra, Martinez Alexander |

| | | | |
|---|---|---|---|
| EPI_ISL_1509002 | Institut National d'hygiène | Unité Mixte Internationale TransVIHMI (UMI 233 IRD - U1175 INSERM - Université de Montpellier) IRD (Institut de recherche pour le développement) | Mounerou SALOU, Christelle BUTEL, Wembo A. HALATOKO,Amivi EHLAN, Abla A. KONOU, Issaka Maman,Syntyche DEVATCHAGNI, Adodo SADJI, Kokou TEGUENI,Koku AGBODEKA, Sidonie A.M.KAGNISSODE, Akoélé SILIADIN, Alassane OURO-MEDELI, Messanh DOUFFAN,Déléma MABA,Sika DOSSIM, Améyo DORKENOO, Mireille PRINCE-DAVID,Anoumou DAGNRA, Laetitia SERRANO,Ahidjo AYOUBA, Eric DELAPORTE, Martine PEETERS |
| EPI_ISL_1510212 | Life Sciences Center, Vilnius University | Vilnius University Hospital Santaros Klinikos, Center of Laboratory Medicine | Gytis Dudas, Ingrida Olendraite, Rimvydas Norvilas, Daniel Naumovas, Dovile Ezerskyte, Ligita Raugaite, Monika Katenaite, Mindaugas Stoskus, Laimonas Griskevicius |
| EPI_ISL_1516300 | OK Public Health Laboratory, Oklahoma State DOH | Centers for Disease Control and Prevention Division of Viral Diseases, Pathogen Discovery | Mili Sheth, Sarah Nobles, Jasmine Padilla, Mark Burroughs, Shoshona Le, Katie Dillon, Peter Cook, Clinton R. Paden, Dhwani Batra, Krista Queen, Kristen Knipe, Dakota Howard, Yvette Unoarumhi, Darlene Wagner, Matthew Schmerer, Ben L. Rambo-Martin, Kristine Lacek, Sam Shepard, Alison Laufer Halpin, Dave Wentworth, Vivien Dugan, Suxiang Tong, Justin Lee |
| EPI_ISL_1517423 | Inciensa, Instituto Costarricense de Investigación y Enseñanza en Nutrición y Salud | Inciensa, Instituto Costarricense de Investigación y Enseñanza en Nutrición y Salud | Cristian Peréz-Corrales, Valeria Peralta-Barquero & Gallegos-Carrillo B |
| EPI_ISL_1522216 | Dutch COVID-19 response team | National Institute for Public Health and the Environment (RIVM) | Adam Meijer, Harry Vennema, Dirk Eggink, Jeroen Cremer, Sharon van den Brink, Bas van der Veer, AnneMarie van den Brandt, Lisa Wijsman, Kim Freriks, Ryanne Jaarsma, Euníce Then, Jolienke Hardeman, Lynn Aarts, Sanne Bos, Melissa van Tuil, Robert Kohl, Linda van de Nes, Sjoerd Kuiling, James Groot, Florian Zwagemaker, Dennis Schmitz, Annelies Kroneman, Karim Hajji, Chantal Reusken, on behalf of the national COVID-19 response team |
| EPI_ISL_1533610 | Shiraz University | Shiraz University | Abozar Ghorbani, Hakimeh Ahmadi, Hamed Gouklani, Ebrahim Eftekhar |
| EPI_ISL_1533989 | Laboratorio Nacional de Salud | Laboratory of Respiratory Viruses and Measles, Oswaldo Cruz Institute, FIOCRUZ | Paola Resende, Cesar Roberto Conde Pereira, Claudia Estrada, Luciana Appolinario, Fernando Motta, Anna Carolina Paixao, Ana Carolina Mendonca, Marilda Siqueira on behalf of the Fiocruz COVID-19 Genomic Surveillance Network |
| EPI_ISL_1540680, EPI_ISL_1540683 | NMVRVI | Lithuanian University of Health Sciences Hospital, Department of Genetics and Molecular Medicine | Rasa Ugenskiene, Darius Cereskevicius, Inga Nasvytiene, Zivile Zemeckiene, Mantas Sarauskas, Marius Sukys, Rima Vainoriene, Astra Vitkauskiene, Renaldas Jurkevicius |
| EPI_ISL_1543465, EPI_ISL_1543808 | Pandemic Response Lab - NYC | Pandemic Response Lab, R&D | Henry Lee, Michael Hammerling, Melissa Hopkins, Cybill del Castillo, Shinyoung Clair Kang, William Ward, Pradeep Bugga, Sol Rey, Dylan Law, Katharine Nelson, Haiping Hao, Jon Laurent |
| EPI_ISL_1545316, EPI_ISL_1545320 | Instituto Nacional de Investigação em Saúde | KRISP, KZN Research Innovation and Sequencing Platform | Morais J, Neto Z, Afonso P, Miranda J, David K, Inglês L, Pereira A, Paulo A Carralero RR Paixão JP, Freitas RH, Mufinda M, Lutucuta S, Giandhari J, Pillay S, Naidoo Y, Emmanuel SJ, Tegally H, Wilkinson E, de Oliveira T |
| EPI_ISL_1560167 | Aegis Sciences Corporation | Centers for Disease Control and Prevention Division of Viral Diseases, Pathogen Discovery | Dakota Howard, Dhwani Batra, Peter W. Cook, Kara Moser, Adrian Paskey, Jason Caravas, Benjamin Rambo-Martin, Shatavia Morrison, Christopher Gulvick, Scott Sammons, Yvette Unoarumhi, Darlene Wagner, Matthew Schmerer, Cyndi Clark, Patrick Campbell, Rob Case, Vikramsinha Ghorpade, Holly Houdeshell, Ola Kvalvaag, Dillon Nall, Ethan Sanders, Alec Vest, Shaun Westlund, Matthew Hardison, Clinton R. Paden, Duncan MacCannell |
| EPI_ISL_1565237 | amedes MVZ Hannover | Robert Koch Institute | unknown |
| EPI_ISL_1566470, EPI_ISL_1566566, EPI_ISL_1568272 | Synlab MVZ Augsburg | Robert Koch Institute | unknown |
| EPI_ISL_1571290 | Eurofins LifeCodexx GmbH | Robert Koch Institute | unknown |
| EPI_ISL_1575038 | Alaska State Virology Laboratory | Alaska State Virology Laboratory | Stephanie DeRonde, Elva House, Lisa Smith, Ph.D., Jack Chen, Ph.D. |
| EPI_ISL_1576771 | MD PHL | MD PHL | Maryland Department of Health Laboratories Administration |
| EPI_ISL_1576950 | UAB Medicina practica laboratorija | Lithuanian University of Health Sciences | Arnoldas Pautienius, Kamile Tamusauskaite, Gediminas Alzbutas, Dovydas Gecys, Lukas Zemaitis, Vaiva Lesauskaite |
| EPI_ISL_1577798 | INHRR | Laboratorio de Virología Molecular | Loureiro CL, Jaspe RC, D Angelo P, Zambrano JL, Rodriguez L, Alarcon V, Delgado M, Aguilar M, Garzaro D, Rangel HR, Pujol FH |
| EPI_ISL_1579527, EPI_ISL_1579571, EPI_ISL_1579607 | NMVRVI | National Public Health Surveillance Laboratory | Lukas Zemaitis, Migle Gabrielaite, Jelena Razmuk, Svajune Muralyte, Ana Steponkiene, Lukas Vasionis, Danas Baksa |
| EPI_ISL_1579691 | NVSPL | National Public Health Surveillance Laboratory | Lukas Zemaitis, Migle Gabrielaite, Jelena Razmuk, Svajune Muralyte, Ana Steponkiene, Lukas Vasionis, Danas Baksa |
| EPI_ISL_1579784, EPI_ISL_1579903 | NMVRVI | National Public Health Surveillance Laboratory | Lukas Zemaitis, Migle Gabrielaite, Jelena Razmuk, Svajune Muralyte, Ana Steponkiene, Lukas Vasionis, Danas Baksa |
| EPI_ISL_1582678 | Hospital | National Reference Center for Viruses of Respiratory Infections, Institut Pasteur, Paris | Marion Barbet, Sylvie Behillil, Frédéric Lemoine, Corinne Maufrais, Christophe Malabat,Gael Millot, Méline Bizard, Angela Brisebarre, Camille Capel, Louise Lefrançois, Etienne Simon-Lorière, Vincent Enouf, Maud Vanpeene, Sylvie van der Werf,Pascale Martres |
| EPI_ISL_1583188 | Armed Forces Institute of Pathology (AFIP), Dhaka Cantonment | Genomic Research Lab, BCSIR | Md. Murshed Hasan Sarkar, Abu Sayeed Mohammad Mahmud, Mohammad Samir Uzzaman, Eshrar Osman, Md. Ahasan Habib, Shahina Akter, Tanjina Akhter Banu, Barna Goswami, Iffat Jahan, Md. Saddam Hossain, Mohammad Mohi Uddin, Md. Kamrul Islam, Mohammad Mizanur Rahman, Susane Giti, Md. Salim Khan |
| EPI_ISL_1583577, EPI_ISL_1583618 | Microbiology Department, Laboratori Clínic Metropolitana Nord. Hospital Universitari Germans Trias i Pujol. | Can Ruti SARS-CoV-2 Sequencing Hub (HUGTiP/IrsiCaixa/IGTP) | Marc Noguera-Julian, Pilar Armengol, Ignacio Blanco, Antoni E Bordoy, Francesc Catala-Moll, Pere-Joan Cardona, Maria Casadellà, Cristina Casañ, Gemma Clara, Bonaventura Clotet, Cristina Esteban, Montserrat Giménez, Mercedes Guerrero, Anna Not, Roger Paredes, Mariona Parera, Verónica Saludes, Alba Sánchez, and Elisa Martró on behalf of the Can Ruti SARS-CoV-2 Sequencing Hub. |
| EPI_ISL_1585395 | Centro de Investigación Biomédica del Noreste (CIBIN) | Instituto Nacional de Enfermedades Respiratorias (INER): Centro de Investigación en Enfermedades Infecciosas (CIENI) | Consorcio Mexicano de Vigilancia Genómica (CoViGen-Mex). Authors (in alphabetical order): Julio Elias Alvarado-Yaah, Carlos F. Arias, Santiago Ávila-Ríos, Víctor Hugo Borja-Aburto, Celia Boukadida, Juan Bautista Chale-Dzul, Célida Duque Molina, José Antonio Enciso-Moreno, Gloria Elena Espinosa-Ayala, Fernando Fontove-Herrera, Victor Eduardo Garcia-Arias, Concepción Grajales-Muñiz, Ricardo Grande, Alfredo Herrera-Estrella, Carla Ivón Herrera-Najera, Pavel Isa, Brenda Irasema Maldonado-Meza, Bernardo Martínez-Miguel, Margarita Matías-Florentino, María Guadalupe de Jesús Mireles-Rivera, Gloria María Molina-Salinas, Hector Montoya-Fuentes, José Esteban Muñoz-Medina, José de Jesús Nuñez-Contreras, Alicia Ocaña-Mondragón, Luis Alberto Ochoa-Carrera, Hector Esteban Paz-Juárez, Francisco Pulido, Helen Haydee Fernanda Ramírez-Plascencia, Angel Gustavo Salas-Lais, Alejandro Sanchez-Flores, Clara Esperanza Santacruz-Tinoco, María Guadalupe Santiago-Mauricio, Nelly Sélem-Mojica, Blanca Taboada, Gloria Vazquez |
| EPI_ISL_1585832, EPI_ISL_1585833 | Vilnius University Hospital Santaros Klinikos | Vilnius University Hospital Santaros Klinikos, Center of Laboratory Medicine | Dovile Ezerskyte, Daniel Naumovas, Gytis Dudas, Ingrida Olendraite, Rimvydas Norvilas, Ligita Raugaite, Monika Katenaite, Mindaugas Stoskus, Laimonas Griskevicius |
| EPI_ISL_1587703 | Infinity Biologix | Centers for Disease Control and Prevention Division of Viral Diseases, Pathogen Discovery | Dakota Howard, Dhwani Batra, Peter W. Cook, Kara Moser, Adrian Paskey, Jason Caravas, Benjamin Rambo-Martin, Shatavia Morrison, Christopher Gulvick, Scott Sammons, Yvette Unoarumhi, Darlene Wagner, Matthew Schmerer, Christian Bixby, Yihe Wang, Jonathan Schultz, Chirayu Goswami, Russ Hager, Robin Grimwood, Clinton R. Paden, Duncan MacCannell |
| EPI_ISL_1590223 | Centrl Laboratorija; Eurofins Genomics Europe Sequencing GmbH | Riga East University Hospital-National Microbiology Reference Laboratory; Eurofins Genomics Europe Sequencing GmbH | irts Šenders,Reinis Vangravs,Arzu Algulieva,Reinis Zeltmatis,Drta Ppola,Ilva Pole,Dina Dusacka,Sergejs Nikisins,Stella Lapia,Jana Oste |
| EPI_ISL_1591299 | Teaching Institute for Public Health of Split-Dalmatia County | Croatian Institute of Public Health | Irena Tabain, Ivana Ferenak |
| EPI_ISL_1595851 | Genome Analysis Center, Yamanashi Central Hospital | Genome Analysis Center, Yamanashi Central Hospital | Yosuke Hirotsu |
| EPI_ISL_1608021 | Jessa | Jessa | Cruys et al. on behalf of the Jessa_cmdLab |
| EPI_ISL_1620228 | OLVZ Aalst | OLVZ Aalst | Astrid Holderbeke |
| EPI_ISL_1623167, EPI_ISL_1623586 | SELARL MIRIALIS CLUSES | CNR Virus des Infections Respiratoires - France SUD | Antonin Bal, Gregory Destras, Gwendolyne Burfin, Hadrien Regue, Quentin Semanas, Martine Valette, Bruno Lina, Laurence Josset |
| EPI_ISL_1624880 | Berkeley Medical Center | WVU and Marshall University Combined Genomics Core Facilities | James Denvir, Peter Stoilov, Peter Perrotta, Wesley Kimble, Ryan Percifield |
| EPI_ISL_1643902, EPI_ISL_1643915 | Eurofins LifeCodexx GmbH | Robert Koch Institute | unknown |
| EPI_ISL_1656854, EPI_ISL_1656882 | NVSPL | National Public Health Surveillance Laboratory | Lukas Zemaitis, Ingrida Olendraite, Arnoldas Pautienius, Kamile Tamusauskaite, Dovydas Gecys, Laura Pareckaite, Vaiva Lesauskaite, Astra Vitkauskiene |

| | | | |
|---|---|---|---|
| EPI_ISL_1661639 | UAB Medicina practica laboratorija | Vilnius University Hospital Santaros Klinikos, Center of Laboratory Medicine | Gytis Dudas, Daniel Naumovas, Dovile Ezerskyte, Ingrida Olendraite, Rimvydas Norvilas, Ligita Raugaite, Monika Katenaite, Mindaugas Stoskus, Laimonas Griskevicius |
| EPI_ISL_1661662 | Vilnius University Hospital Santaros Klinikos | Vilnius University Hospital Santaros Klinikos, Center of Laboratory Medicine | Gytis Dudas, Daniel Naumovas, Dovile Ezerskyte, Ingrida Olendraite, Rimvydas Norvilas, Ligita Raugaite, Monika Katenaite, Mindaugas Stoskus, Laimonas Griskevicius |
| EPI_ISL_1661694, EPI_ISL_1661705, EPI_ISL_1661716, EPI_ISL_1661727, EPI_ISL_1661738 | Anteja laboratorija (UAB Diagnostikos laboratorija) | Vilnius University Hospital Santaros Klinikos, Center of Laboratory Medicine | Gytis Dudas, Daniel Naumovas, Dovile Ezerskyte, Ingrida Olendraite, Rimvydas Norvilas, Ligita Raugaite, Monika Katenaite, Mindaugas Stoskus, Laimonas Griskevicius |
| EPI_ISL_1661747 | Vilnius University Hospital Santaros Klinikos | Vilnius University Hospital Santaros Klinikos, Center of Laboratory Medicine | Gytis Dudas, Daniel Naumovas, Dovile Ezerskyte, Ingrida Olendraite, Rimvydas Norvilas, Ligita Raugaite, Monika Katenaite, Mindaugas Stoskus, Laimonas Griskevicius |
| EPI_ISL_1671822 | CH Roubaix | CHU Lille - Laboratoire de Virologie | AIT YAHYA Emilie, ALIDJINOU Enagnon Kazali, BOCKET Laurence, CREPIN Michel, DEMAY Christophe, ENGELMANN Ilka, GEFFROY Sandrine, GUIGON Aurélie, LAMBERT Valérie, LAZREK Mouna, NOBILLIAUX Florian, PREVOST Brigitte, THUILLIER Caroline, TINEZ Claire |
| EPI_ISL_1673323 | Laboratorio de Investigaciones de Baney | Swiss Tropical and Public Health Institute | Salome Hosch, Carlos Cortes, Claudia Daubenberger, Guillermo Garcia, Bonifacio Manguire Nlavo, Maximilian Mpina, Elizabeth Nyakarungu, Diosdado Odjama Nseng Ada, Mitoha Ondo O Ayekaba, Tobias Schindler, Philipp Wagner, Philip Wonder Phiri |
| EPI_ISL_1675013 | Labo Analyses Med | National Reference Center for Viruses of Respiratory Infections, Institut Pasteur, Paris | Marion Barbet, Sylvie Behillil, Méline Bizard, Angela Brisebarre, Camille Capel, Vincent Enouf, Louise Lefrançois, Frédéric Lemoine, Christophe Malabat, Corinne Maufrais, Pierre Lechat, Etienne Simon-Lorière, Maud Vanpeene, Sylvie Van der Werf ,Karine Breant |
| EPI_ISL_1675021 | Labo Analyses Med | National Reference Center for Viruses of Respiratory Infections, Institut Pasteur, Paris | Marion Barbet, Sylvie Behillil, Méline Bizard, Angela Brisebarre, Camille Capel, Vincent Enouf, Louise Lefrançois, Frédéric Lemoine, Christophe Malabat, Corinne Maufrais, Pierre Lechat, Etienne Simon-Lorière, Maud Vanpeene, Sylvie Van der Werf ,Fabienne Artur |
| EPI_ISL_1675082, EPI_ISL_1675084 | Hospital | National Reference Center for Viruses of Respiratory Infections, Institut Pasteur, Paris | Marion Barbet, Sylvie Behillil, Méline Bizard, Angela Brisebarre, Camille Capel, Vincent Enouf, Louise Lefrançois, Frédéric Lemoine, Christophe Malabat, Corinne Maufrais, Etienne Simon-Lorière, Maud Vanpeene, Sylvie Van der Werf ,Pascale Martres |
| EPI_ISL_1675656 | Institute of Molecular and Translational Medicine / Laboratory of Experimental Medicine, Faculty of Medicine and Dentistry, Palacky University and University Hospital Olomouc | Institute of Molecular and Translational Medicine / Laboratory of Experimental Medicine, Faculty of Medicine and Dentistry, Palacky University | Rastislav Slavkovský, Hana Jaworek, Vladimíra Koudeláková, Barbora Blumová, Tomáš Pospíšil, Marián Hajdúch |
| EPI_ISL_1688635, EPI_ISL_1688660 | Lab voor klinische biologie | Lab voor klinische biologie | Marija Janevska, Hannelore Hamerlinck, Bruno Verhasselt |
| EPI_ISL_1700651 | Hospital | National Reference Center for Viruses of Respiratory Infections, Institut Pasteur, Paris | Marion Barbet, Sylvie Behillil, Méline Bizard, Angela Brisebarre, Camille Capel, Vincent Enouf, Louise Lefrançois, Frédéric Lemoine, Christophe Malabat, Corinne Maufrais, Adrien Pain, Etienne Simon-Lorière, Maud Vanpeene, Sylvie Van der Werf, Pascale Martres |
| EPI_ISL_1715187, EPI_ISL_1715188, EPI_ISL_1715190, EPI_ISL_1715191, EPI_ISL_1715192, EPI_ISL_1715193, EPI_ISL_1715194, EPI_ISL_1715195, EPI_ISL_1715196, EPI_ISL_1715197, EPI_ISL_1715199, EPI_ISL_1715201, EPI_ISL_1715203 | | | |
| see above | National Public Health Laboratory, Cameroon | African Centre of Excellence for Genomics of Infectious Diseases, Redeemer's University | Olawoye, I.B. et al |
| EPI_ISL_1718408 | Lighthouse Lab in Alderley Park | Wellcome Sanger Institute for the COVID-19 Genomics UK (COG-UK) Consortium | Jacquelyn Wynn, Mairead Hyland, The Lighthouse Lab in Alderley Park and Alex Alderton, Roberto Amato, Jeffrey Barrett, Sonia Goncalves, Ewan Harrison, David K. Jackson, Ian Johnston, Dominic Kwiatkowski, Cordelia Langford, John Sillitoe on behalf of the Wellcome Sanger Institute COVID-19 Surveillance Team |
| EPI_ISL_1750667 | Viollier AG | Department of Biosystems Science and Engineering, ETH Zürich | Chaoran Chen, Sarah Nadeau, Ivan Topolsky, Emmanouil Dermitzakis, Keith Harshman, Ioannis Xenarios, Henri Pegeot, Lorenzo Cerutti, Deborah Penet, Philipp Jablonski, Lara Fuhrmann, David Dreifuss, Katharina Jahn, Christiane Beckmann, Maurice Redondo, Olivier Kobel, Christoph Noppen, Sophie Seidel, Noemie Santamaria de Souza, Niko Beerenwinkel, Tanja Stadler |
| EPI_ISL_1755103 | Hôpital Necker-Enfants malades | Department of Virology, Henri Mondor University Hospital, Assistance Publique Hôpitaux de Paris, Université Paris-Est Créteil, INSERM U955 | Christophe Rodriguez, Slim Fourati, Vanessa Demontant, Guillaume Gricourt, Melissa N'Debi, Alexandre Soulier, Elisabeth Trawinski, Jean-Michel Pawlotsky |
| EPI_ISL_1756769 | UAB Medicina practica laboratorija | Vilnius University Hospital Santaros Klinikos, Center of Laboratory Medicine | Ingrida Olendraite, Gytis Dudas, Daniel Naumovas, Dovile Ezerskyte, Rimvydas Norvilas, Ligita Raugaite, Monika Katenaite, Mindaugas Stoskus, Laimonas Griskevicius |
| EPI_ISL_1756904 | UAB InMedica | Vilnius University Hospital Santaros Klinikos, Center of Laboratory Medicine | Ingrida Olendraite, Gytis Dudas, Daniel Naumovas, Dovile Ezerskyte, Rimvydas Norvilas, Ligita Raugaite, Monika Katenaite, Mindaugas Stoskus, Laimonas Griskevicius |
| EPI_ISL_1757704 | CH.INTERCOMMUNAL DE CRETEIL | Department of Virology, Henri Mondor University Hospital, Assistance Publique Hôpitaux de Paris, Université Paris-Est Créteil, INSERM U955 | Christophe Rodriguez, Slim Fourati, Vanessa Demontant, Guillaume Gricourt, Melissa N'Debi, Alexandre Soulier, Elisabeth Trawinski, Jean-Michel Pawlotsky |
| EPI_ISL_1757775 | Hôpital Necker-Enfants malades | Department of Virology, Henri Mondor University Hospital, Assistance Publique Hôpitaux de Paris, Université Paris-Est Créteil, INSERM U955 | Christophe Rodriguez, Slim Fourati, Vanessa Demontant, Guillaume Gricourt, Melissa N'Debi, Alexandre Soulier, Elisabeth Trawinski, Jean-Michel Pawlotsky |
| EPI_ISL_1760006, EPI_ISL_1760007, EPI_ISL_1760008, EPI_ISL_1760009, EPI_ISL_1760020, EPI_ISL_1760021, EPI_ISL_1760022, EPI_ISL_1760023 | Life Sciences Center, Vilnius University | Institute of Biotechnology, Life Sciences Center, Vilnius University | Emilija Vasiliunaite, Milda Norkiene, Danguole Ziogiene, Albertas Timinskas, Alma Gedvilaite |
| EPI_ISL_1761494, EPI_ISL_1761505 | Microbiology Department, Laboratori Clínic Metropolitana Nord. Hospital Universitari Germans Trias i Pujol. | Can Ruti SARS-CoV-2 Sequencing Hub (HUGTiP/IrsiCaixa/IGTP) | Marc Noguera-Julian, Pilar Armengol, Ignacio Blanco, Antoni E Bordoy, Francesc Catala-Moll, Pere-Joan Cardona, Maria Casadellà, Cristina Casañ, Gemma Clara, Bonaventura Clotet, Cristina Esteban, Montserrat Giménez, Mercedes Guerrero, Anna Not, Roger Paredes, Mariona Parera, Verónica Saludes, Alba Sánchez, and Elisa Martró on behalf of the Can Ruti SARS-CoV-2 Sequencing Hub. |
| EPI_ISL_1785368 | National Virus Reference Laboratory | National Virus Reference Laboratory | Fiona Crispie, Calum Walsh, Matthew McCabe, Zoe Yandle, Charlene Bennet, Gabriel Gonzalez, Michael Carr, Jonathan Dean, Paul Cotter, Cillian F De Gascun |
| EPI_ISL_1789037 | Hospital | National Reference Center for Viruses of Respiratory Infections, Institut Pasteur, Paris | Marion Barbet, Sylvie Behillil, Méline Bizard, Angela Brisebarre, Camille Capel, Vincent Enouf, Louise Lefrançois, Frédéric Lemoine, Christophe Malabat, Corinne Maufrais, Etienne Simon-Lorière, Maud Vanpeene, Sylvie Van der Werf ,CéLine Bressollette |
| EPI_ISL_1789089, EPI_ISL_1789090, EPI_ISL_1789091, EPI_ISL_1789092, EPI_ISL_1789093, EPI_ISL_1789094, EPI_ISL_1789095, EPI_ISL_1789096, EPI_ISL_1789097 | Hospital | National Reference Center for Viruses of Respiratory Infections, Institut Pasteur, Paris | Marion Barbet, Sylvie Behillil, Méline Bizard, Angela Brisebarre, Camille Capel, Vincent Enouf, Louise Lefrançois, Frédéric Lemoine, Christophe Malabat, Corinne Maufrais, Etienne Simon-Lorière, Maud Vanpeene, Sylvie Van der Werf ,Pascale Martres |
| EPI_ISL_1789100 | Hospital | National Reference Center for Viruses of Respiratory Infections, Institut Pasteur, Paris | Marion Barbet, Sylvie Behillil, Méline Bizard, Angela Brisebarre, Camille Capel, Vincent Enouf, Louise Lefrançois, Frédéric Lemoine, Christophe Malabat, Corinne Maufrais, Etienne Simon-Lorière, Maud Vanpeene, Sylvie Van der Werf ,JéRome Guinard |
| EPI_ISL_1805832 | Johns Hopkins Hospital Department of Pathology | Johns Hopkins Hospital Department of Pathology | C. Paul Morris, Chun Huai Luo, Adannaya Amadi, Matthew Schwartz, Heba H. Mostafa |
| EPI_ISL_1817033 | MEPHI, Aix Marseille University | MEPHI, Aix Marseille University | Anthony LEVASSEUR |
| EPI_ISL_1821604 | Hospital | National Reference Center for Viruses of Respiratory Infections, Institut Pasteur, Paris | Marion Barbet, Sylvie Behillil, Méline Bizard, Angela Brisebarre, Camille Capel, Vincent Enouf, Louise Lefrançois, Frédéric Lemoine, Christophe Malabat, Corinne Maufrais, Damien Mornico, Etienne Simon-Lorière, Maud Vanpeene, Sylvie Van der Werf ,Pascale Martres |
| EPI_ISL_1823196 | Vilnius University Hospital Santaros Klinikos | Vilnius University Hospital Santaros Klinikos, Center of Laboratory Medicine | Ingrida Olendraite, Gytis Dudas, Daniel Naumovas, Dovile Ezerskyte, Rimvydas Norvilas, Ligita Raugaite, Monika Katenaite, Mindaugas Stoskus, Laimonas Griskevicius |
| EPI_ISL_1823197, EPI_ISL_1823198, | UAB InMedica | Vilnius University Hospital Santaros Klinikos, Center of | Ingrida Olendraite, Gytis Dudas, Daniel Naumovas, Dovile Ezerskyte, Rimvydas Norvilas, Ligita Raugaite, Monika Katenaite, Mindaugas Stoskus, |

| | | | |
|---|---|---|---|
| EPI_ISL_1823199 | | Laboratory Medicine | Laimonas Griskevicius |
| EPI_ISL_1823200 | Vilnius University Hospital Santaros Klinikos | Vilnius University Hospital Santaros Klinikos, Center of Laboratory Medicine | Ingrida Olendraite, Gytis Dudas, Daniel Naumovas, Dovile Ezerskyte, Rimvydas Norvilas, Ligita Raugaite, Monika Katenaite, Mindaugas Stoskus, Laimonas Griskevicius |
| EPI_ISL_1823201 | SYNLAB Lietuva UAB | Vilnius University Hospital Santaros Klinikos, Center of Laboratory Medicine | Ingrida Olendraite, Gytis Dudas, Daniel Naumovas, Dovile Ezerskyte, Rimvydas Norvilas, Ligita Raugaite, Monika Katenaite, Mindaugas Stoskus, Laimonas Griskevicius |
| EPI_ISL_1827069, EPI_ISL_1827401 | Lab voor klinische biologie | Lab voor klinische biologie | Marija Janevska, Hannelore Hamerlinck, Bruno Verhasselt |
| EPI_ISL_1828718 | National Institute of Public Health | State Veterinary Institute Prague | Nagy,A;Jirincova,H;Suri,T;Trnka,D;Vecerova,J |
| EPI_ISL_1829052, EPI_ISL_1829167 | CHUV | Laboratory of genomics and metagenomics | Trestan Pillonel, Damien Jacot, Sébastien Aeby, Gilbert Greub, Claire Bertelli |
| EPI_ISL_1843903 | Centogene; Dr. Bauer Laboratoriums GmbH | Robert Koch Institute | unknown |
| EPI_ISL_1844130, EPI_ISL_1844259 | Sonic - MVZ Medizinisches Labor Bremen GmbH | Robert Koch Institute | unknown |
| EPI_ISL_1845707 | SYNLAB MVZ Weiden | Robert Koch Institute | unknown |
| EPI_ISL_1845845, EPI_ISL_1845921, EPI_ISL_1845926 | Eurofins LifeCodexx GmbH | Robert Koch Institute | unknown |
| EPI_ISL_1847168, EPI_ISL_1847176 | MVZ Labor Dr. Limbach & Kollegen GbR | Robert Koch Institute | unknown |
| EPI_ISL_1852817 | SYNLAB MVZ Weiden | Robert Koch Institute | unknown |
| EPI_ISL_1854262 | Instituto Nacional de Saude (INSA) and Institute of Biomedicine (iBiMed), Universidade de Aveiro | Instituto Nacional de Saude (INSA) and Institute of Biomedicine (iBiMed), Universidade de Aveiro | Borges et al |
| EPI_ISL_402125 | National Institute for Communicable Disease Control and Prevention (ICDC) Chinese Center for Disease Control and Prevention (China CDC) | National Institute for Communicable Disease Control and Prevention (ICDC) Chinese Center for Disease Control and Prevention (China CDC) | Zhang,Y.-Z., Wu,F., Chen,Y.-M., Pei,Y.-Y., Xu,L., Wang,W., Zhao,S., Yu,B., Hu,Y., Tao,Z.-W., Song,Z.-G., Tian,J.-H., Zhang,Y.-L., Liu,Y., Zheng,J.-J., Dai,F.-H., Wang,Q.-M., She,J.-L. and Zhu,T.-Y. |
| EPI_ISL_406798 | General Hospital of Central Theater Command of People's Liberation Army of China | BGI & Institute of Microbiology, Chinese Academy of Sciences & Shandong First Medical University & Shandong Academy of Medical Sciences & General Hospital of Central Theater Command of People's Liberation Army of China | Weijun Chen, Yuhai Bi, Weifeng Shi and Zhenhong Hu |
| EPI_ISL_416538 | Wellington Hospital | Institute of Environmental Science and Research (ESR) | Wellington SCL, Wellington Hospital, Riddiford Street, Newtown, Wellington 6021, New Zealand |
| EPI_ISL_426539 | AZ SPHL, Arizona Department of Health Services | TGen North | Jolene Bowers, Megan Folkerts, Darrin Lemmer, Dave Engelthaler |
| EPI_ISL_437451 | B.J. Medical College and Civil hospital | Gujarat Biotechnology Research Centre | Kairavi Joshi, Gaurishankar Shrimali, Nidhi Sood, Pranay Shah, R D Dixit, Snehal Bagatharia, Kamlesh J Upadhyay, Ramesh Pandit, Tejas Shah, Ankit Hinsu, Pritesh Sabara, Apurvasinh Puvar, Janvi Raval, Monika Gandhi, Pinal Trivedi, Maharshi Pandya, Amit Kanani, Akanksha Verma, Nitin Savaliya, Raghawendra Kumar, Dinesh Kumar, Zuber Saiyed, Dipa Kinariwala, Disha Patel, Binita Aring, Neeta Khandelwal, Geeta Vaghela, Sonia Barve, Bhavesh Modi, Bhavya Jindal, Chaitanya Joshi, Madhvi Joshi |
| EPI_ISL_444999 | Naval Health Research Center | Naval Medical Research Center Biological Defense Research Directorate | Logan Voegtly, Regina Cer, Dessiree Pena-Gomez, Adrian Paskey,Kyle Long, Roger Pan, Melinda Balansay-Ames, Chris Myers, Ewell Hollis, Nathaniel Christy, Kimberly Bishop-Lilly |
| EPI_ISL_455077 | South Eastern Area Laboratory Services | NSW Health Pathology - Institute of Clinical Pathology and Medical Research; Westmead Hospital; University of Sydney | CIDM-PH et al. |
| EPI_ISL_456382 | LabPLUS | Institute of Environmental Science and Research (ESR) | Matt Storey, Xiaoyun Ren, Anja Werno, Antje van der Linden, Arlo Upton, Chris Mansell, David Hammer, Dragana Drinkovic, Erasmus Smit, Gary McAuliffe, Hana Sofia Andersson, James Ussher, Jill Sherwood, Josh Freeman, Julia Howard, Juliet Elvy, Mary DeAlmeida, Matt Blakiston, Matthew Rogers, Max Bloomfield, Michael Addidle, Michelle Balm, Sally Roberts, Sarah Jefferies, Sharmini Muttaiyah, Susan Morpeth, Susan Taylor, Timothy Blackmore, Vani Sathyendran, Veronica Playle, Virginia Hope, Erasmus Smit, Lauren Jelly, Joep de Ligt |
| EPI_ISL_469254 | National Institute for Viral Disease Control and Prevention, China CDC | Institute of Viral Disease Control and Prevention, China CDC | Wenjie Tan, Lijuan Chen, Peihua NiuBaoying Huang, Li Zhao, Yubai Bi, Wenling Wang, Roujian Lu, Dayan Wang, Wenbo Xu, George Fu Gao, Chun Huang, Guizhen Wu |
| EPI_ISL_479581 | National Public Health Laboratory, National Centre for Infectious Diseases | National Public Health Laboratory, National Centre for Infectious Diseases | Mak TM, Octavia S, Zhou Z, Chavatte JM, Cui L, Lin RTP |
| EPI_ISL_482959 | Minnesota Department of Health, Public Health Laboratory | Minnesota Department of Health, Public Health Laboratory | Matt Plumb, Jacob Garfin, and Xiong Wang |
| EPI_ISL_491951 | Instituto Nacional de Investigación en Salud Pública - INSPI | INSPI - Charité | Alfredo Bruno Caicedo, Domenica de Mora Coloma, Andres Moreira-Soto, Anna-Lena Sander, Nina Krause, Maritza Olmedo,Denisses Portugal, Manuel Gonzalez, Silvia Salgado, Alberto Orlando, Alexandra Usiña, Juan Carlos Zeballos, Jan Felix Drexler |
| EPI_ISL_492020 | Oman-NIC | Department of Microbiology and Immunology-SQUH | Fahad Zadjali, Samira Al-Maruqi, Amina Al Jardani, Khulood Al-Mammary, Hanan Al-kindi, Fatma BaAlawi, Hamida AL Barwani, Zeyana AL-Dahmani, Intisar Al-Shukri, Aisha Al-Busaidi, Aisha Al-Amri, Ahlam Al-Amri, Mohammed Al-Tobi, Samiha Al Kharusi, Abdulla Balkhair |
| EPI_ISL_496790 | Gorgas Memorial Laboratory of Health Studies | Gorgas Memorial Laboratory of Health Studies | Danilo Franco, Claudia Gonzalez Sandra Lopez-Verges, Alexander A Martinez |
| EPI_ISL_498694 | National Institute for Viral Disease Control and Prevention, China CDC | National Institute for Viral Disease Control and Prevention, China CDC | Xiang Zhao,LingLing Mao,Yao Meng,Zhixiao Chen,Yuchao Wu,Yong ZhangBo ZhijianJianqun Zhang,Yang Song,Dayan Wang,WenQing YaoWenbo Xu |
| EPI_ISL_527877 | Nigeria Centre for Disease Control (NCDC) | African Centre of Excellence for Genomics of Infectious Diseases (ACEGID), Redeemer's University, Ede, Osun State, Nigeria | Oluniyi P.E. et al |
| EPI_ISL_529213 | Beijing Institute of Microbiology and Epidemiology | Beijing Institute of Microbiology and Epidemiology | Fan, Hang; Qin, E.; Wu, Y.; Guo, Y.; Zhang, X.; Yong, Y.; Hou, J.; Xu, Z.; Mu, J.; Teng, Yue; Mi, Z.; Yang, R.; Song, Yajun.; Li, B.; Cui, Y. |
| EPI_ISL_529742 | NHLS-IALCH | KRISP, KZN Research Innovation and Sequencing Platform | Giandhari J, Pillay S, Lessells R, Mdlalose K, York D, Khan S, Tegally H, Wilkinson E, de Oliveira T |
| EPI_ISL_530250 | Queensland Health Forensic and Scientific Services, Public Health Virology | Public Health Virology Laboratory, Forensic and Scientific Services, Queensland Health | Son Nguyen et al |
| EPI_ISL_536504 | Instituto Nacional de Salud | Laboratorio de Infecciones Respiratorias Agudas | Eduardo Juscamayta Lopez, David Tarazona, Faviola Valdivia Guerrero, Nancy Rojas Serrano, Dennis Carhuaricra, Lenin Maturrano Hernandez, Ronnie Gavilan Chavez |
| EPI_ISL_548145 | Middlemore Hospital | Institute of Environmental Science and Research (ESR) | Xiaoyun Ren, Matt Storey, Nikki Freed, Muhammad Faisal, Jing Wang, Hermes Perez, Anja Werno, Antje van der Linden, Arlo Upton, Chris Mansell, David Hammer, Dragana Drinkovic, Gary McAuliffe, Hana Sofia Andersson, James Ussher, Jill Sherwood, Josh Freeman, Julia Howard, Juliet Elvy, Mary DeAlmeida, Matt Blakiston, Matthew Rogers, Max Bloomfield, Michael Addidle, Michelle Balm, Sally Roberts, Sarah Jefferies, Sharmini Muttaiyah, Susan Morpeth, Susan Taylor, Timothy Blackmore, Vani Sathyendran, Veronica Playle, Virginia Hope, Erasmus Smit, Lauren Jelly, Olin Silander, Joep de Ligt |
| EPI_ISL_572163 | Quest Diagnostics | Quest Diagnostics | Rosenthal,S.H., Gerasimova,A., Kagan,R.M., Anderson, B., Grover, D., Livingston, K.E., Hua, M., Liu Y., Shalhout, D.F., Owen, R., Lacbawan, F. |
| EPI_ISL_577741 | Institute of Virology, Biomedical Research Center of the Slovak Academy of Sciences, Bratislava | Faculty of Natural Sciences, Comenius University, Bratislava | Broa Brejová, Viktória Hodorová, Kristína Boršová, Viktória abanová, Dominika Friová, Sabina Fumaová Havlíková, Juraj Kopáek, Martina Liková, ubomíra Lukáiková, Martina Neboháová, Monika Sláviková, Edita Staroová, Elena Tichá, Tomáš Vina, Jozef Nosek, Boris Klempa |
| EPI_ISL_579406 | North Shore Hospital | Institute of Environmental Science and Research (ESR) | Xiaoyun Ren, Matt Storey, Nikki Freed, Muhammad Faisal, Jing Wang, Hermes Perez, Anja Werno, Antje van der Linden, Arlo Upton, Chris Mansell, David Hammer, Dragana Drinkovic, Gary McAuliffe, Hana Sofia Andersson, James Ussher, Jill Sherwood, Josh Freeman, Julia Howard, Juliet Elvy, Mary DeAlmeida, Matt Blakiston, Matthew Rogers, Max Bloomfield, Michael Addidle, Michelle Balm, Sally Roberts, Sarah Jefferies, Sharmini Muttaiyah, Susan Morpeth, Susan Taylor, Timothy Blackmore, Vani Sathyendran, Veronica Playle, Virginia Hope, Erasmus Smit, Lauren Jelly, Olin Silander, Joep de Ligt |

| | | | |
|---|---|---|---|
| EPI_ISL_583896 | Singapore General Hospital | Department of Microbiology | Nurdyana Abdul Rahman, Kun Lee Lim, Chenhao Li, Sui Sin Goh, Kenneth Xin Long Chan, Kian Sing Chan, Lynette Oon, Kern Rei Chng, Niranjan Nagarajan, Karrie Ko |
| EPI_ISL_590728 | University of Michigan Clinical Microbiology Laboratory | Lauring Lab, University of Michigan, Department of Microbiology and Immunology | Valesano |
| EPI_ISL_591277 | National Institute for Viral Disease Control and Prevention, China CDC | National Institute for Viral Disease Control and Prevention, China CDC | Huilai Ma, Zhaoguo Wang, Xiang Zhao, Jun Han, Yong Zhang, Hong Wang, Cao Chen, Ji Wang, Jingdong Song, Yao Meng, Yuchao Wu, Zhixiao Chen, Dayan Wang, Ruqin Gao, George F.Gao, Wenbo Xu |
| EPI_ISL_601443 | Lighthouse Lab in Milton Keynes | Wellcome Sanger Institute for the COVID-19 Genomics UK (COG-UK) consortium | The Lighthouse Lab in Milton Keynes and Alex Alderton, Roberto Amato, Sonia Goncalves, Ewan Harrison, David K. Jackson, Ian Johnston, Dominic Kwiatkowski, Cordelia Langford, John Sillitoe on behalf of the Wellcome Sanger Institute COVID-19 Surveillance Team (http://www.sanger.ac.uk/covid-team) |
| EPI_ISL_632284 | Communicable Disease Laboratory, Public Health Directorate | Communicable Disease Laboratory, Public Health Directorate | AlWasti,H., AlTaif,Z., AlHujairi,Z., AlAbbas,Z. |
| EPI_ISL_636980 | CS Xai Xai | KRISP, KZN Research Innovation and Sequencing Platform | Ismael N, Giandhari J, Pillay S, Tegally H, Wilkinson E, de Oliveira T, Nadia Sitoe, Paulo Arnaldo, Nedio Mabunda |
| EPI_ISL_648672 | Department of Laboratory Medicine, Tan Tock Seng Hospital | Department of Laboratory Medicine, Tan Tock Seng Hospital | Chen YYC, Zair X, Lim JX, Li C, Tang WY, Maurer-Stroh S, Barkham TMS, Nagarajan N, Sessions OM |
| EPI_ISL_660464 | Laboratoire de Microbiologie CHU Souro Sanou | Centre Muraz | Abdoul-Salam Ouedraogo, Yacouba Sawadogo, Essia Belarbi, Grit Schubert, Fabian Leendertz, Arsène Zongo, Soumeya Ouangraoua, Zekiba Tarnagda, Lassana Sangaré, Halidou Tinto |
| EPI_ISL_671437 | University of Debrecen, Department of Medical Microbiology | National Laboratory of Virology, Szentágothai Research Centre | Endre Gábor Tóth, Balázs Somogyi, Brigitta Zana, Eszter Csoma, Ferenc Jakab, Gábor Kemenesi |
| EPI_ISL_693831 | Viollier AG | Department of Biosystems Science and Engineering, ETH Zürich | Christian Beisel, Sarah Nadeau, Chaoran Chen, Ivan Topolsky, Pedro Ferreira, Philipp Jablonski, Susana Posada-Céspedes, Tobias Schär, Ina Nissen, Natascha Santacroce, Elodie Burcklen, Christiane Beckmann, Maurice Redondo, Olivier Kobel, Christoph Noppen, Sophie Seidel, Noemie Santamaria de Souza, Niko Beerenwinkel, Tanja Stadler |
| EPI_ISL_697791 | Institute of Microbiology, Universidad San Francisco de Quito | Institute of Microbiology, Universidad San Francisco de Quito | Belén Prado-Vivar, Sully Márquez, Juan José Guadalupe, Monica Becerra-Wong, Bernardo Gutiérrez, Jonathan Araujo, Verónica Barragán, Patricio Rojas-Silva, Gabriel Trueba, Michelle Grunauer, Paúl Cárdenas |
| EPI_ISL_708815 | Urban Institute for Disease Prevention and Control | National Institute of Health, Department of Medical Sciences, Ministry of Public Health, Thailand | Pilailuk Okada; Siripaporn Phuygun; Thanutsapa Thanadachakul; Sittiporn Parnmen; Pakorn Piromtong; Warawan Wongboot; Sunthareeya Waicharoen; Malinee Chittaganpitch |
| EPI_ISL_710570 | University Hospital Dubrava | Ruer Boškovic Institute; Forensic Science Centre Ivan Vueti; University of Zagreb Faculty of Science | Robert Beluži, Marina Korolija, Ana Livun, Vjekoslav Tomai, Dunja Glavaš, Maja Kuzman, Paula Štancl, Lucija Markulin, Lucija Basi, Antonela Blažekovi, Fran Boroveki, Lidija Cvetko-Krajinovi, Ivana elap, Fuad osovi, Mirjana Domazet-Lošo, Tomislav Domazet-Lošo, Valentina umlijan-Combaj, Kristina Gotovac Jerei, Jasna Kašman, Vladimir Krajinovi, Danilo Licastro, Boris Maek, Željka Maak Šafranko, Gordana Maravi Vlahoviek, Senica Pejša, Josipa Skelin, Ivan Šamija, Mario Štefanovi, Sanja Tadinac, Katarina Marija Tupek, Petra Vrabec, Rosa Karli, Kristian Vlahoviek |
| EPI_ISL_712081 | Port Elizabeth Provincial Hospital, National Health Laboratory Services, Eastern Cape, South Africa | National Institute for Communicable Diseases of the National Health Laboratory Service | Mohale T, Ntuli N, Mahlangu B, Allam M, Ismail A, Bhiman JN |
| EPI_ISL_718146 | Ministry of Health Hospitals | Institute of Health and Community Medicine | David Perera, Ooi Mong How, Chua Hock Hin, Tonnii Sia Loong Loong, Wong Jyn Shan, Wong Kiing Aik, Chan Chia Jui |
| EPI_ISL_728204 | Institute of Microbiology, Universidad San Francisco de Quito | Institute of Microbiology, Universidad San Francisco de Quito | Sully Márquez, Belén Prado-Vivar, Juan José Guadalupe, Monica Becerra-Wong, Bernardo Gutiérrez, Tania Guayasamin, Patricio Reyes, Verónica Barragán, Patricio Rojas-Silva, Gabriel Trueba, Michelle Grunauer, Paúl Cárdenas |
| EPI_ISL_732954 | Department of Tropical Parasitology | Laboratory of Recombinant Vaccines | Lukasz Rabalski, Maciej Kosinski, Teemu Smura, Kirsi Aaltonen, Ravi Kant, Tarja Sironen, Boguslaw Szewczyk, Maciej Grzybek |
| EPI_ISL_740868 | South Eastern Area Laboratory Services (SEALS) | NSW Health Pathology - Institute of Clinical Pathology and Medical Research; Westmead Hospital; University of Sydney | CIDM-PH et al. |
| EPI_ISL_746484 | Genetica Molecular and Subdepartamento de Virologia ISP Chile | Instituto de Salud Publica de Chile | Javier Tognarelli, Barbara Parra, Loredana Arata, Jaime Lagos, Gisselle Barra, Patricia Bustos, Rodrigo Fasce, Andres Castillo, Jorge Fernandez |
| EPI_ISL_749906 | Sanatorio Americano | Institut Pasteur de Montevideo | Daiana Mir, Natalia Rego, Paola Cristina Resende, Fernando Lopez-Tort, Tamara Fernandez-Calero, Veronica Noya, Mariana Brandes, Tania Possi, Mailen Arleo, Natalia Reyes, Matias Victoria, Andres Lizasoain, Matias Castells, Leticia Maya, Matías Salvo, Tatiana Schäffer Gregianini, Marilda Tereza Mar da Rosa, Letícia Garay Martins, Cecilia Alonso, Yasser Vega, Cecilia Salazar, Ignacio Ferrés, Pablo Smircich, Jose Sotelo, Ighor Arantes, Luciana Appolinario, Ana Carolina Mendonça, Maria Jose Benitez-Galeano, Martín Graña, Camila Simoes, Fernando Motta, Marilda Mendonça Siqueira, Gonzalo Bello, Rodney Colina, Lucia Spangenberg |
| EPI_ISL_766025 | Department of Virology and Immunology, University of Helsinki and Helsinki University Hospital, Huslab Finland | Department of Virology, Faculty of Medicine, University of Helsinki, Helsinki, Finland | Teemu Smura, Olli Vapalahti, Maija Lappalainen, Satu Kurkela |
| EPI_ISL_767874 | Histopath | NSW Health Pathology - Institute of Clinical Pathology and Medical Research; Westmead Hospital; University of Sydney | CIDM-PH et al. |
| EPI_ISL_779409 | Royal Darwin Hospital Pathology | MDU-PHL | Meumann, E., Caly L., Seemann T., Sait, M.L., Druce J., Sherry, N.L. |
| EPI_ISL_792350 | Laboratorio del Hospital Interzonal General de Agudos Evita | Área de Secuenciación del Laboratorio de Virología del Hospital de Niños Dr. Ricardo Gutierrez on behalf of 'Proyecto Argentino Interinstitucional de genomica de SARS-CoV-2' (PAIS Consortium) | Nabaes Jodar, MS; Goya, S; Natale, MI; Lusso, S; Desimone, I; Luczac, E; Serrano, L; Grossi, O; Musto, A; Valinotto, LE; Viegas, M. |
| EPI_ISL_802863 | Vilnius University Hospital Santaros Klinikos, Vilnius University | Institute of Biotechnology, Life Sciences Center, Vilnius University | Emilija Vasiliunaite, Milda Norkiene, Albertas Timinskas, Alma Gedvilaite, Aurelija Zvirbliene, Daniel Naumovas, Laimonas Griskevicius |
| EPI_ISL_806849 | Alaska State Virology Laboratory | Alaska State Virology Laboratory | Stephanie DeRonde, Lisa Smith, Ph.D., Devin M. Drown, Ph.D., Jack Chen, Ph.D. |
| EPI_ISL_807156 | Deva County Emergency Hospital | National Institute of Infectious Diseases-Prof. Dr. Matei Bals Molecular Diagnostics Laboratory | Leontina Banica, Marius Surleac, Corina Casangiu, Petre Milu, Andreea Tudor, Simona Paraschiv, Dan Otelea |
| EPI_ISL_810801, EPI_ISL_810804 | PathWest Laboratory Medicine WA | PathWest Laboratory Medicine WA Microbial Surveillance Unit | PathWest Laboratory Medicine WA Microbial Surveillance Unit |
| EPI_ISL_823886 | DOHMH Central Harlem | New York City Public Health Laboratory | Jade Wang, et al. |
| EPI_ISL_833137 | Laboratorio de Ecologia de Doencas Transmissiveis na Amazonia, Instituto Leonidas e Maria Deane - Fiocruz Amazonia | Laboratorio de Ecologia de Doencas Transmissiveis na Amazonia, Instituto Leonidas e Maria Deane - Fiocruz Amazonia | Valdinete Nascimento, Victor Souza, André Corado, Fernanda Nascimento, George Silva, Ágatha Costa, Debora Duarte, Karina Pessoa, Matilde Mejía, Luciana Gonçalves, Maria Júlia Brandão, Michele Jesus, Felipe Naveca on behalf of the Fiocruz COVID-19 Genomic Surveillance Network |
| EPI_ISL_845545, EPI_ISL_845546, EPI_ISL_845548, EPI_ISL_845549, EPI_ISL_845550, EPI_ISL_845551, EPI_ISL_845552, EPI_ISL_845553, EPI_ISL_845554, EPI_ISL_845557, EPI_ISL_845558, EPI_ISL_845560, EPI_ISL_845561, EPI_ISL_845562, EPI_ISL_845563, EPI_ISL_845564, EPI_ISL_845565 | | | |
| see above | National Public Health Laboratory, Cameroon | African Centre of Excellence for Genomics of Infectious Diseases (ACEGID), Redeemer's University | Oluniyi P.E. et al |
| EPI_ISL_849747 | Public Health Virology Laboratory, Forensic and Scientific Services (PHV-FSS) | Public Health Virology Laboratory, Forensic and Scientific Services (PHV-FSS) | Son Nguyen et al |
| EPI_ISL_850949, EPI_ISL_850951 | National Institute for Viral Disease Control and Prevention, China CDC | National Institute for Viral Disease Control and Prevention, China CDC | Xiang Zhao, Yenan Feng, Zhixiao Chen, Yao Meng, Yuchao Wu, Yang Song, Ji Wang, Kai Nie, Yong Zhang, Yanhai Wang, Weimin Zhou, Wenjie Tan, Jun Han, Shiwen Wang, Wenbo Xu, Cao Chen, Dayan Wang |
| EPI_ISL_862079 | National Influenza Center, Virology Department | National Influenza Center | K Sadeghi, A Nejati, J Yavarian, NZ Shafiei Jandaghi, V Salimi, F Ajaminejad,N Ghavvami and T Mokhtari Azad |
| EPI_ISL_872606 | Nigeria Centre for Disease Control (NCDC) | African Centre of Excellence for Genomics of Infectious | Oluniyi P.E. et al |

| | | | |
|---|---|---|---|
| | | Diseases (ACEGID), Redeemer's University | |
| EPI_ISL_875538 | Institute of Virology, Biomedical Research Center of the Slovak Academy of Sciences, Bratislava | Faculty of Natural Sciences, Comenius University, Bratislava | Viktória abanová, Kristína Boršová, Broa Brejová, Viktória Hodorová, Sabina Fumaová Havlíková, Juraj Kopáek, Martina Liková, ubomíra Lukáiková, Martina Neboháová, Monika Sláviková, Tomáš Vina, Jozef Nosek, Boris Klempa |
| EPI_ISL_887486 | Instituto Nacional de Saude (INS), Mozambique | KRISP, KZN Research Innovation and Sequencing Platform | Nalia Ismael, Nadia Sitoe, Paulo Arnaldo, Nedio Mabunda, Giandhari J, Pillay S, Tegally H, Wilkinson E, de Oliveira T |
| EPI_ISL_907075 | Department of Biology, University of Basrah | Department of Biology, University of Basrah | Abu-Ali,H.M. and Al-Badran,I.F. |
| EPI_ISL_907108 | Cancer Biology Department, National Cancer Institute | Cancer Biology Department, National Cancer Institute | Zekri,A.N., Sedawy,M.G., Ahmed,O.S., Hafez,M.M., Soliman,H.K., Bahnassy,A.A., Elhosieny,F.W., Gad,A.E., Hamdy,M.S., Soliman,M.S., Soliman,L., Abouelhoda,M. |
| EPI_ISL_912369 | Fondation Congolaise pour la recherche medicale (FCRM), Francine Ntoumi | NGS Competence Center Tuebingen, Institut für Medizinische Mikrobiologie und Hygiene, Universitaetsklinikum Tübingen | Angel Angelov |
| EPI_ISL_918371 | Virology Unit, Institut Pasteur du Cambodge | Virology Unit, Institut Pasteur du Cambodge | Sokhoun Yann, Ly Sovann, Kraing Sidonn, Yi Sengdoeurn, Chin Savuth, Chau Darapheak, Etienne Simon-Loriere, Veasna Duong, Erik A Karlsson |
| EPI_ISL_950607 | Queens Medical Centre, Clinical Microbiology Department / DeepSeq Nottingham | COVID-19 Genomics UK (COG-UK) Consortium | Gemma Clark, Wendy Smith, Manjinder Khakh, Vicki M Fleming, Michelle M Lister, Hannah Howson-Wells, Jonathan Ball, Patrick McClure, Joseph Chappell, Theocharis Tsoleridis, Nadine Holmes, Matthew Carlisle, Christopher Moore, Fei Sang, Johnny Debebe, Victoria Wright, Matthew Loose |
| EPI_ISL_956326 | Laboratory Medicine | Department of Laboratory Medicine, Lin-Kou Chang Gung Memorial Hospital, Taoyuan, Taiwan | Kuo-Chien Tsao, Yu-Nong Gong, Shu-Li Yang, Yi-Chun Liu, Chung-Guei Huang, Mei-Jen Hsiao, Po-Wei Huang, Cheng-Ta Yang, Cheng-Hsun Chiu, Peng-Nien Huang, Kuo-Ming Lee, Guang-Wu Chen, Shin-Ru Shih |
| EPI_ISL_968849, EPI_ISL_968873 | KEMRI-Wellcome Trust Research Programme/KEMRI-CGMR-C Kilifi | KEMRI-Wellcome Trust Research Programme/KEMRI-CGMR-C Kilifi | Githinji et al |
| EPI_ISL_977352 | University of Zambia, School of Veterinary Medicine | UNZAVET and PATH | Mulenga Mwenda-Chimfwembe, Ngonda Saasa, Daniel Bridges |
| EPI_ISL_979271 | Cadham Provincial laboratory | National Microbiology Laboratory (NML) | Anna Majer, Shari Tyson, Grace Seo, Philip Mabon, Elsie Grudeski, Rhiannon Huzarewich, Russell Mandes, Anneliese Landgraff, Jennifer Tanner, Natalie Knox, Morag Graham, Gary Van Domselaar, Paul Van Caeseele, Jared Bullard, David Alexander, Kerry Dust, Nathalie Bastien, Yan Li, Timothy Booth, Darian Hole, Madison Chapel, Kirsten Biggar, CanCOGeN's metadata curation team, Public Health Agency of Canada CanCOGeN team |
| EPI_ISL_981024 | National Public Health Laboratory, National Centre for Infectious Diseases | National Public Health Laboratory, National Centre for Infectious Diseases | Tze Minn Mak, Zhenyang Zhou, Lin Cui, Raymond Tzer Pin Lin |
| EPI_ISL_999032 | Institut Pasteur de Guinée | Institut Pasteur de Dakar | Grayo Solene, Diagne Moussa Moïse, Dia Ndongo, Diallo Amadou, Mbengue Safietou Sankhe, Ndiaye Ndack, Diop Mamadou, Loucoubar Cheikh, Tordo Noel, Faye Ousmane, Sall Amadou Alpha |
